# Serum proteomics reveals *APOE* dependent and independent protein signatures in Alzheimer’s disease

**DOI:** 10.1101/2023.11.08.23298251

**Authors:** Elisabet A. Frick, Valur Emilsson, Thorarinn Jonmundsson, Anna E. Steindorsdottir, Erik C. B. Johnson, Raquel Puerta, Eric B. Dammer, Anantharaman Shantaraman, Amanda Cano, Mercè Boada, Sergi Valero, Pablo García-González, Elias F. Gudmundsson, Alexander Gudjonsson, Joseph J. Loureiro, Anthony P. Orth, Nicholas T. Seyfried, Allan I. Levey, Agustin Ruiz, Thor Aspelund, Lori L. Jennings, Lenore J. Launer, Valborg Gudmundsdottir, Vilmundur Gudnason

## Abstract

The current demand for early intervention, prevention, and treatment of late onset Alzheimer’s disease (LOAD) warrants deeper understanding of the underlying molecular processes which could contribute to biomarker and drug target discovery. Utilizing high-throughput proteomic measurements in serum from a prospective population-based cohort of older adults (n=5,294), we identified 303 unique proteins associated with incident LOAD (median follow-up 12.8 years). Over 40% of these proteins were associated with LOAD *independently* of *APOE-*ε*4* carrier status. These proteins were implicated in neuronal processes and overlapped with protein signatures of LOAD in brain and cerebrospinal fluid. We found 17 proteins which LOAD-association was strongly *dependent* on *APOE-*ε*4* carrier status. Most of them showed consistent associations with LOAD in cerebrospinal fluid and a third had brain-specific gene expression. Remarkably, four proteins in this group (TBCA, ARL2, S100A13 and IRF6) were downregulated by *APOE-*ε*4* yet upregulated as a consequence of LOAD as determined in a bi-directional Mendelian randomization analysis, reflecting a potential response to the disease onset. Accordingly, the direct association of these proteins to LOAD was reversed upon *APOE-*ε*4* genotype adjustment, a finding which we replicate in an external cohort (n=719). Our findings provide an insight into the dysregulated pathways that may lead to the development and early detection of LOAD, including those both independent and dependent on *APOE-*ε*4*. Importantly, many of the LOAD-associated proteins we find in the circulation have been found to be expressed - and have a direct link with AD - in brain tissue. Thus, the proteins identified here, and their upstream modulating pathways, provide a new source of circulating biomarker and therapeutic target candidates for LOAD.

## Introduction

Alzheimer‘s disease (AD) is the most common cause of dementia, accounting for up to 80% of all dementia cases^1^, of which *late onset* Alzheimer‘s disease (LOAD) is most common^2^. As of 2022, approximately 55 million individuals worldwide had dementia, representing 1 out of 9 people aged 65 and over^3^. While promising advances have been made in amyloid-targeting therapeutic options for early-stage LOAD^4,5^, they still have limited benefit and identification of additional risk pathways that can be used for early detection and intervention is highly needed. To meet these demands, a variety of biologically relevant circulating molecules have been broadly associated with LOAD risk. The proteome in particular has the potential to reveal circulating markers of disease-related molecular pathways from different tissues, and studies assessing the circulating proteomic signatures between non-demented older adults and individuals suffering from LOAD have been described^6–17^. Modest sample sizes, low-throughput proteomics and lack of longitudinal measurements have, however, been limiting factors in these studies. However, a recent large-scale longitudinal study identified promising blood-based markers for all-cause incident dementia although it is unknown how specific the results are to LOAD^18^. Information on the global circulating proteomic profile preceding the onset of LOAD, and how well it reflects AD-related processes in brain and CSF, is thus scarce.

Alzheimer’s disease has a considerable genetic component, and both common^19^ and rare risk variants have been identified^20^, of which the strongest effects are conferred by variants in the well-known apolipoprotein E (*APOE*) gene. Approximately 25% of the general population carries the *APOE*-ε*4* variant while it is present in over 50% of AD cases^21,22^. The *APOE*-ε4 allele increases the risk of LOAD by threefold in heterozygous carriers and up to twelvefold in homozygous carriers^23^. Although the link between the ε4 allele and LOAD has been extensively researched, light has yet to be shed on the precise mechanism by which the *APOE* gene affects LOAD onset and/or progression. Importantly, recent large-scale proteogenomic studies have consistently established the *APOE* locus as a protein-regulatory hotspot, regulating levels of hundreds of proteins in both circulation^24–27^ and cerebrospinal fluid (CSF)^28,29^. Yet, it remains unknown to what extent these proteins relate to LOAD and if they can provide new information on the mechanisms through which *APOE*-ε4 mediates it risk. Identifying LOAD-associated circulatory proteins and whether their association is *APOE-*dependent or independent is crucial for the understanding of AD more generally as well as for gaining insight into potential pathways suitable for targeting in personalized treatment.

The current study tests the hypotheses that specific proteomic signatures in the circulation precede LOAD diagnosis and can reflect dysregulated biological pathways in the brain and CSF. Furthermore, we expect that some of these protein signatures may be affected by the *APOE-*ε*4* genotype and can thus provide molecular read-out of pathways directly affected by *APOE-*ε*4*. To address these hypotheses, we used a high throughput aptamer-based platform to characterize 4,137 serum proteins in 5,294 participants of the population-based Age, Gene/Environment Susceptibility-Reykjavik Study (AGES-RS)^30^ to identify protein signatures of incident LOAD (events occurring during follow-up) and prevalent LOAD, taking an unbiased, longitudinal, and cross-sectional approach to the discovery of potential biomarkers for LOAD (Figure 1). Considering *APOE*’s protein-regulatory influence and how it may impact the way that serum-proteins are associated with LOAD, we disentangled the LOAD protein signature into *APOE-*ε*4* dependent and independent components, by identifying proteins whose LOAD-association is largely attenuated upon conditioning on *APOE-*ε*4* carrier status. We compared the serum protein signature of LOAD to those observed in cerebrospinal fluid (CSF) and brain and finally, used genetic variation as anchors to determine the potential causal direction between serum proteins and disease state.

**Fig. 1.**
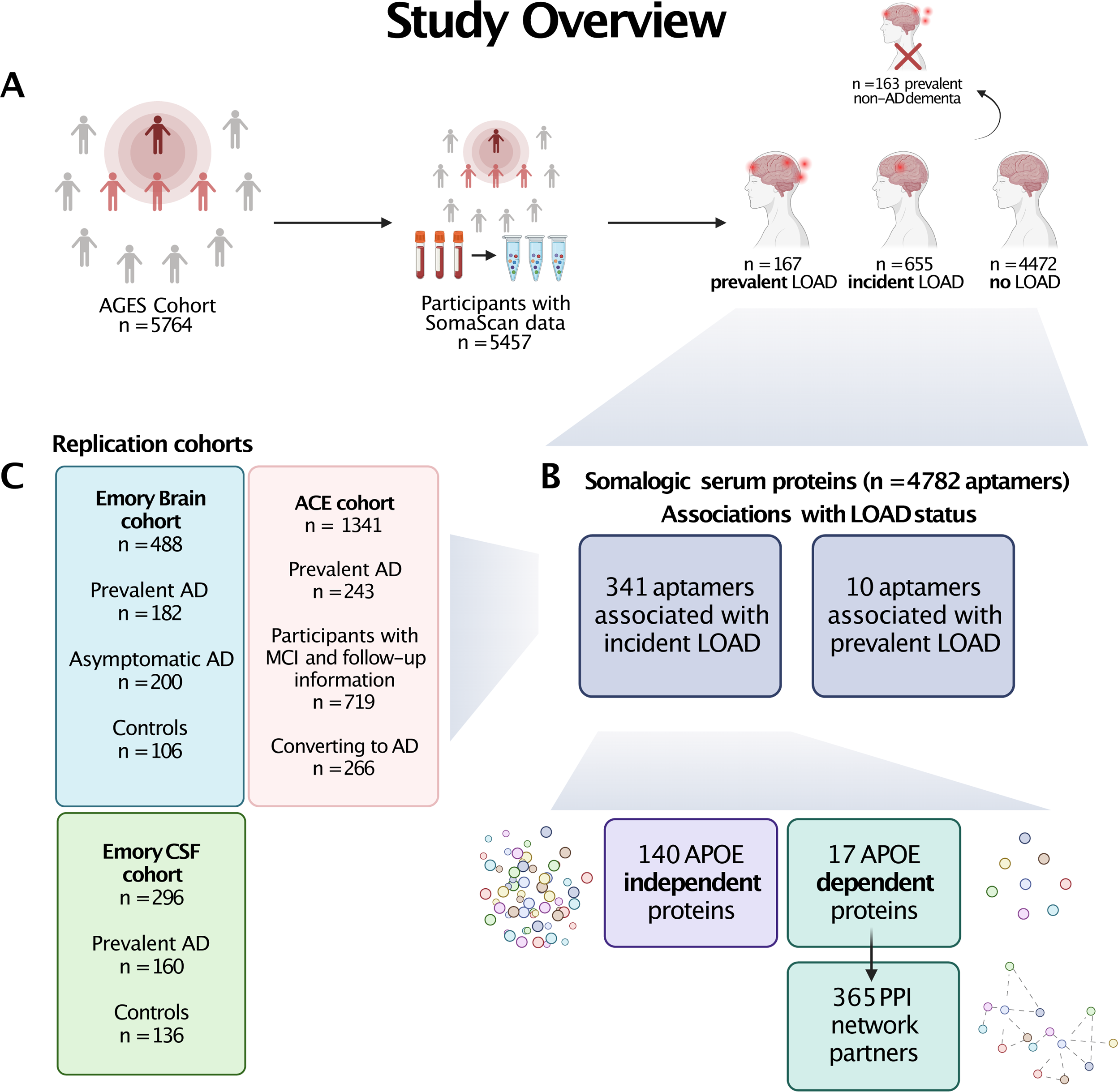
Study overview. Flowchart of the current study. **A)** Overview of the AGES cohort and study participants. Prevalent non-AD dementia cases were excluded from the analysis. **B)** Overview of the aptamers tested and their associations with LOAD. Serum measurements of 4782 aptamers were associated to prevalent and incident LOAD status, using logistic and Cox proportional hazards regression models, respectively. From the proteins associated with incident LOAD, sets of 140 proteins with an *APOE*-independent associations and 17 proteins with an *APOE*-dependent association were defined. The APOE-dependent proteins were further expanded to first degree protein-protein interaction (PPI) partners. All sets of proteins were subjected to functional enrichment analysis and bi-directional Mendelian Randomization (MR) analysis. **C)** Overview of the replication cohorts used in the study which include proteins measured in the circulation (ACE) as well as in brain and CSF (Emory).

## Results

### The AGES study cohor

This prospective population-based study was based on 5,127 participants free of dementia at baseline, after the exclusion of 163 individuals with prevalent non-AD dementia, and 167 individuals with prevalent LOAD. During a potential follow-up of 12.8 years (median), 655 individuals were diagnosed with incident LOAD, with the last individual being diagnosed 16 years from baseline. Participants with incident LOAD were older at entry, were more likely to carry an *APOE-*ε*4* allele, had lower BMI, and had lower education levels compared to healthy individuals (Supplementary Table 1). See Figure 1 for study overview.

### Serum protein profile of incident LOAD in AGES

To investigate the LOAD-associated circulatory proteomic patterns which occur prior to disease onset, we used Cox proportional hazards models and found 320 aptamers (303 proteins) to be significantly (FDR < 0.05) associated with incident LOAD diagnosis after adjusting for age and sex (model 1), with hazard ratios (HRs) ranging from 0.78 for TBCA to 1.47 for NTN1 (Figure 2D, Supplementary Table 2). To account for variability related to *APOE-*ε*4* carrier status, we adjusted for the genotype in an additional model (model 2, Supplementary Table 2), which resulted in 140 significant aptamers (130 proteins, HR 0.79 (CD4) – 1.25 (CGA/FSHB), FDR < 0.05) (Figure 2E), all of which overlapped with model 1 (Figure 2F). When comparing the two models, 43% of the serum proteins remained significant after *APOE-*ε*4* adjustment, indicating that their LOAD association is independent of the *APOE-*ε*4* genotype (Table 1). Adjusting for additional AD risk factors and eGFR (see Methods) retained 38 significant LOAD-associated aptamers (35 proteins, HR 0.80 (CD4) – 1.26 (SMOC1), FDR < 0.05) (model 3, Supplementary Table 2), which may reflect specific processes affecting risk of LOAD but not driven by currently established risk factors.

**Fig. 2.**
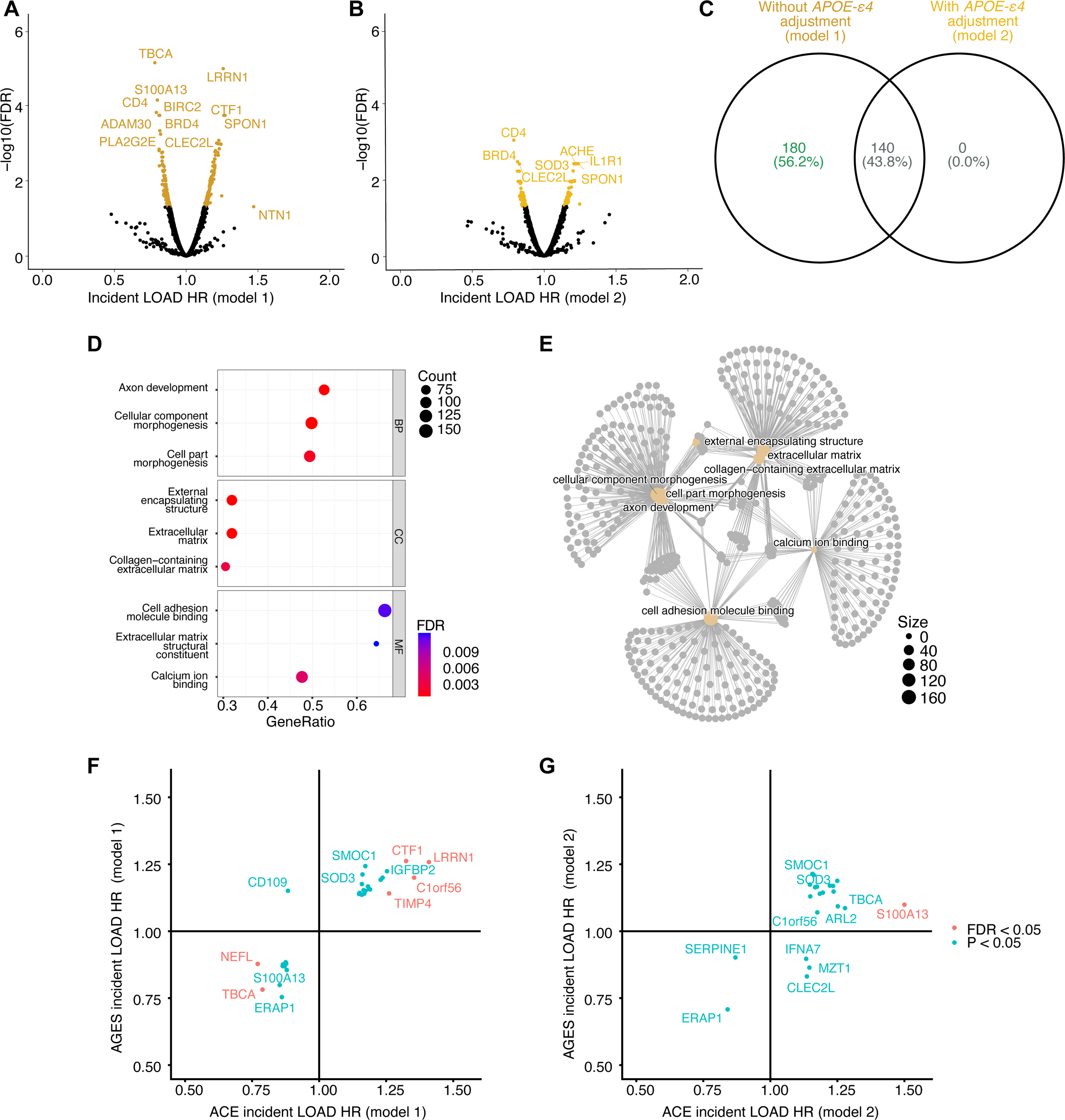
Proteins associated with LOAD in AGES. **A-B)** Volcano plots showing the protein association profile for incident LOAD **A)** without *APOE-*ε*4* adjustment (model 1) and **B)** with *APOE-*ε*4* adjustment (model 2). **C)** Venn diagram for the overlap between models 1 and 2 for incident LOAD. **D-E)** Enrichment of top Gene Ontology terms from GSEA analysis for incident LOAD (model 1) shown as **D)** dotplot, stratified by ontology and **E)** gene-concept network. **F-G)** Comparison of effect sizes (HR) for incident LOAD between the AGES and the ACE cohorts for all proteins reaching nominal significance (P < 0.05) in ACE for **F)** model 1 and **G)** model 2.

**Table 1.**
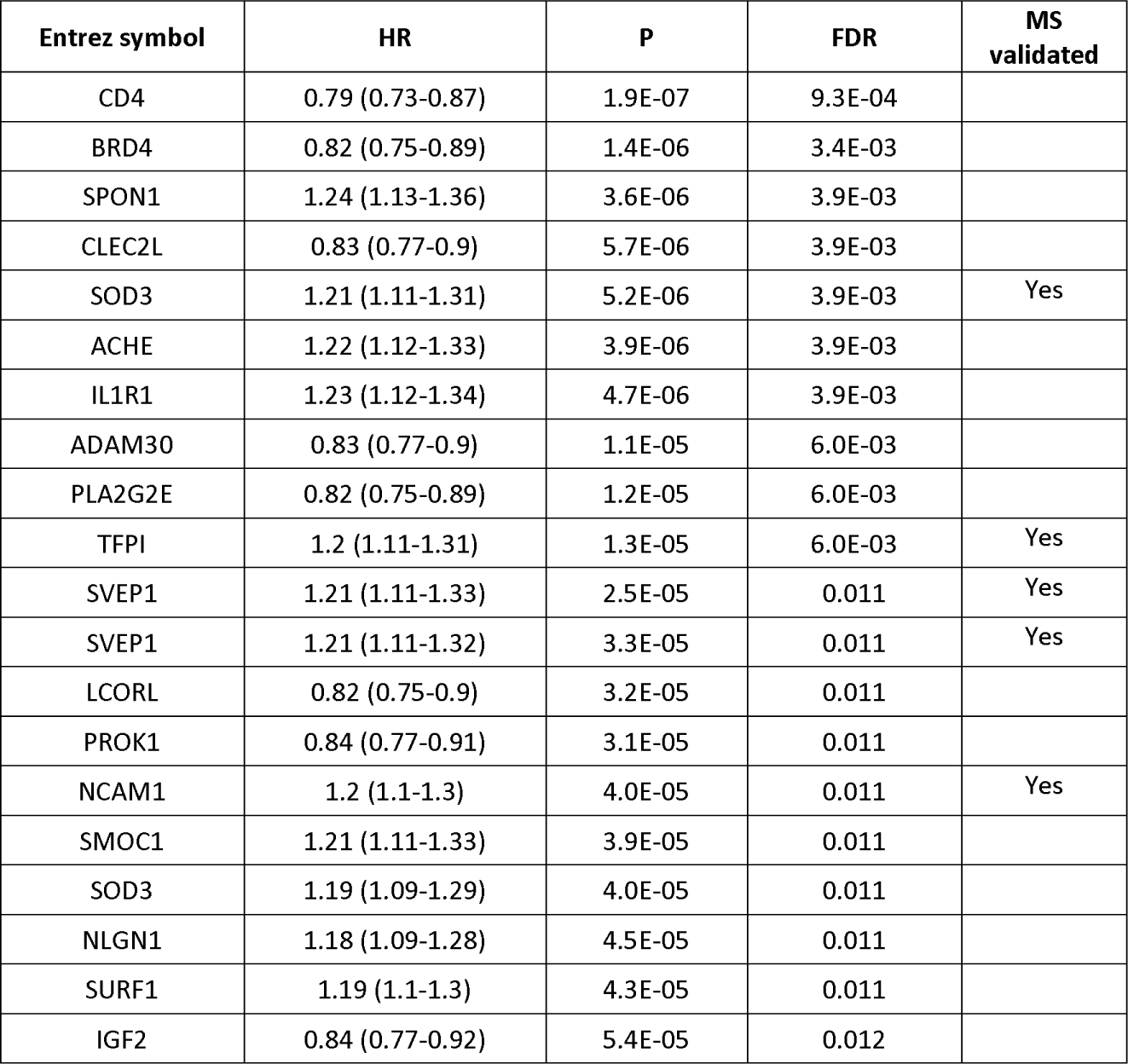
A summary table of the top 20 significant *APOE-independent* proteins associated with incident LOAD in AGES. The effect size (Hazard Ratio (Confidence Interval)) and level of significance (P, FDR) is shown for model 2, adjusting for age, sex and *APOE-ε4*. The final column indicates if the aptamers have been orthogonally validated by mass-spectrometry^26^. The *APOE*-independency is defined as proteins remaining significantly (FDR < 0.05) associated with incident LOAD after *APOE-ε4* adjustment.

| Entrez symbol | HR | P | FDR | MS validated |
| --- | --- | --- | --- | --- |
| CD4 | 0.79 (0.73-0.87) | 1.9E-07 | 9.3E-04 |  |
| BRD4 | 0.82 (0.75-0.89) | 1.4E-06 | 3.4E-03 |  |
| SPON1 | 1.24 (1.13-1.36) | 3.6E-06 | 3.9E-03 |  |
| CLEC2L | 0.83 (0.77-0.9) | 5.7E-06 | 3.9E-03 |  |
| SOD3 | 1.21 (1.11-1.31) | 5.2E-06 | 3.9E-03 | Yes |
| ACHE | 1.22 (1.12-1.33) | 3.9E-06 | 3.9E-03 |  |
| IL1R1 | 1.23 (1.12-1.34) | 4.7E-06 | 3.9E-03 |  |
| ADAM30 | 0.83 (0.77-0.9) | 1.1E-05 | 6.0E-03 |  |
| PLA2G2E | 0.82 (0.75-0.89) | 1.2E-05 | 6.0E-03 |  |
| TFPI | 1.2 (1.11-1.31) | 1.3E-05 | 6.0E-03 | Yes |
| SVEP1 | 1.21 (1.11-1.33) | 2.5E-05 | 0.011 | Yes |
| SVEP1 | 1.21 (1.11-1.32) | 3.3E-05 | 0.011 | Yes |
| LCORL | 0.82 (0.75-0.9) | 3.2E-05 | 0.011 |  |
| PROK1 | 0.84 (0.77-0.91) | 3.1E-05 | 0.011 |  |
| NCAM1 | 1.2 (1.1-1.3) | 4.0E-05 | 0.011 | Yes |
| SMOC1 | 1.21 (1.11-1.33) | 3.9E-05 | 0.011 |  |
| SOD3 | 1.19 (1.09-1.29) | 4.0E-05 | 0.011 |  |
| NLGN1 | 1.18 (1.09-1.28) | 4.5E-05 | 0.011 |  |
| SURF1 | 1.19 (1.1-1.3) | 4.3E-05 | 0.011 |  |
| IGF2 | 0.84 (0.77-0.92) | 5.4E-05 | 0.012 |  |

As hazard ratio variability can arise with lengthy follow-up time, secondary analyses were implemented with a 10-year follow-up cut-off, which revealed mostly overlapping results (Supplementary Note 1, Supplementary Tables 3 and 4, Supplementary Figure 1). We did however detect protein associations specific to the shorter follow-up time, which potentially reflect processes that take place closer to the LOAD diagnosis. As there may be further differences in proteomic profiles depending on whether protein sampling occurred before or after LOAD diagnosis, we additionally considered the protein profile of the 167 individuals with prevalent LOAD at baseline (Supplementary Note 2, Supplementary Figure 2A-C, Supplementary Tables 5-7). Interestingly, many of the proteins associated with increased risk of incident LOAD showed the opposite direction of effect for prevalent LOAD, although generally not statistically significant (Supplementary Figure 2D). These contrasting results suggest an important temporal element in the LOAD-associated proteome. In total there were 346 aptamers (329 unique proteins) associated with LOAD when all outcomes (incident and prevalent LOAD), follow-up times and models were considered (Supplementary Tables 2, 3, 5, 6, and 7).

To evaluate which biological processes are reflected by the overall incident LOAD-associated protein signature in AGES, we performed a gene set enrichment analysis (GSEA). The strongest enrichment for protein associations in model 1 was observed for gene ontology (GO) terms related to neuron development and morphogenesis (Figure 2H-I, Supplementary Table 8). The proteins driving the enrichment included Neural Cell Adhesion Molecules 1 and 2 (NCAM1, and NCAM2), Netrin 1 (NTN1), Contactin 1 (CNTN1), Neuropilin 1 (NRP1), Fibronectin Leucine Rich Transmembrane Protein 2 (FLRT2), Matrix Metallopeptidase 2 (MMP2) and Cell Adhesion Molecule L1 like (CHL1). GSEA of the protein profiles of model 2, where *APOE-*ε*4* carrier status was adjusted for, showed similar enrichment results (Supplementary Table 8), demonstrating that these terms were mainly driven by the *APOE-*ε*4* independent component of the LOAD-associated protein profile.

### Serum proteins with APOE-dependent association to incident LOAD

As previously mentioned, 43% of the protein associations with incident LOAD were independent of *APOE-*ε*4*. Of the remaining 57% that were affected by *APOE-*ε*4* adjustment, we identified 17 proteins whose associations with incident LOAD were particularly strongly affected by *APOE-*ε*4* carrier status (Table 2, Figure 3A, Supplementary Figure 3, Supplementary Table 2). These proteins, hereafter referred to as *APOE*-dependent proteins, were defined as proteins significantly (FDR < 0.05) associated with incident LOAD in *model 1* but whose nominal significance was attenuated (P > 0.05) or direction of effect changed upon *APOE-*ε*4* adjustment in *model 2*. These *APOE*-dependent proteins included those with the strongest associations to LOAD prior to adjusting for the *APOE-*ε*4* allele (Figure 2A, D). The levels of the APOE protein itself were not associated with either incident or prevalent LOAD. Figure 3B shows the intra-correlations among the 17 *APOE*-dependent proteins. All the 17 *APOE-*dependent proteins were strongly regulated by the *APOE-*ε*4* allele (Figure 3C, Table 2, Supplementary Figure 4, Supplementary Table 9), with the ε*4* allele increasing the levels of five of the proteins and decreasing the levels of the other 12. Accordingly, we observed that *increased* levels of the five *APOE-*ε*4* upregulated proteins and *decreased* levels of the 12 *APOE-*ε*4* downregulated proteins were also associated with higher risk of LOAD, yielding a hazard ratio above and below one, respectively (Figure 3D). As per definition, most of the *APOE-*dependent proteins lost significance upon *APOE-*ε*4* adjustment yet interestingly, the direction of effect inverted for five proteins after *APOE-*ε*4* adjustment (ARL2, IRF6, NEFL, S100A13, TBCA) (Figure 3E).

**Table 2.** A summary table of the 17 *APOE*-dependent LOAD associated proteins, describing their tissue specificity in the Human Protein Atlas v22, results from the association analyses in AGES, references for previous associations with *APOE* or LOAD and whether the aptamers have been orthogonally validated via MS ^26^. The *APOE*-dependency is defined as being significant (FDR<0.05) in model 1 and fully non-significant in model 2 (P>0.05) for incident LOAD.

|  |  |  |  |  | Incident LOAD (Cox) |  |  |  | APOE-e4 (linear regression) |  |  |  |  |
| --- | --- | --- | --- | --- | --- | --- | --- | --- | --- | --- | --- | --- | --- |
|  |  |  |  |  | Model 1 |  | Model 2 |  |  |  |  |  |  |
| Entrez Symbol | Tissue specificity | Tissue cluster | LOAD ref | APOE ref | HR | FDR | HR | P | Beta | SE | P | FDR | MS validation |
| LRRN1 | Brain | Brain | 59 | 49 | 1.258 | 1.E-05 | 1.018 | 0.711 | 1.061 | 0.023 | <1E-300 | <1E-300 |  |
| S100A13 | Low | Mitochondria |  | 49 | 0.799 | 7.E-05 | 1.099 | 0.092 | -1.090 | 0.021 | <1E-300 | <1E-300 |  |
| TBCA | Low | Vesicular transport |  | 49 | 0.782 | 7.E-06 | 1.086 | 0.145 | -1.183 | 0.020 | <1E-300 | <1E-300 | Yes |
| CTF1 | Low | Fibroblasts, ECM | 60 | 49 | 1.262 | 2.E-04 | 1.077 | 0.147 | 0.611 | 0.021 | 1.E-171 | 5.E-171 |  |
| ARL2 | Low | Striated muscle |  |  | 0.835 | 0.002 | 1.093 | 0.077 | -0.991 | 0.022 | <1E-300 | <1E-300 |  |
| C1orf56 | Testis | Spermatogenesis |  |  | 1.200 | 0.002 | 1.070 | 0.113 | 0.513 | 0.026 | 6.E-85 | 9.E-85 |  |
| MSN | Low | Neutrophils, Inflammatory response | 61 |  | 1.186 | 0.003 | 1.039 | 0.388 | 0.608 | 0.025 | 9.E-122 | 2.E-121 |  |
| TMCC3 | Brain | Brain, Nervous system development | 62 |  | 0.849 | 0.005 | 0.962 | 0.364 | -0.517 | 0.026 | 4.E-86 | 7.E-86 |  |
| HBQ1 | Bone marrow | Neutrophils, Humoral immune response |  |  | 0.860 | 0.013 | 0.966 | 0.423 | -0.455 | 0.025 | 1.E-69 | 1.E-69 |  |
| IRF6 | Esophagus, Skin | Squamous epithelial cells |  |  | 0.876 | 0.032 | 1.033 | 0.458 | -0.675 | 0.025 | 4.E-153 | 1.E-152 |  |
| IFIT2 | Bone marrow | Brain & bone marrow, Chromatin organization |  |  | 0.875 | 0.035 | 0.949 | 0.214 | -0.275 | 0.026 | 2.E-26 | 2.E-26 |  |
| TP53I11 | Low | Adipose tissue, ECM organization |  |  | 0.879 | 0.036 | 0.981 | 0.653 | -0.436 | 0.026 | 1.E-60 | 1.E-60 |  |
| FAM159B | Brain and other | Unknown function |  |  | 0.882 | 0.039 | 0.977 | 0.591 | -0.419 | 0.026 | 8.E-56 | 9.E-56 |  |
| NDE1 | Low | Brain, Nervous system development | 63 |  | 0.876 | 0.039 | 1.001 | 0.985 | -0.489 | 0.025 | 3.E-80 | 5.E-80 | Yes |
| NEFL | Brain | Brain, Neuronal signaling | 64 |  | 0.878 | 0.041 | 1.031 | 0.516 | -0.578 | 0.024 | 2.E-121 | 4.E-121 |  |
| GSTM1 | Brain | Liver, Metabolism | 65 | 66 | 1.136 | 0.042 | 1.077 | 0.087 | 0.276 | 0.026 | 1.E-26 | 1.E-26 |  |
| GGT2 | Kidney, Thyroid | Kidney, Transmembrane transport |  |  | 0.887 | 0.048 | 0.981 | 0.646 | -0.384 | 0.026 | 2.E-47 | 3.E-47 |  |

**Fig. 3.**
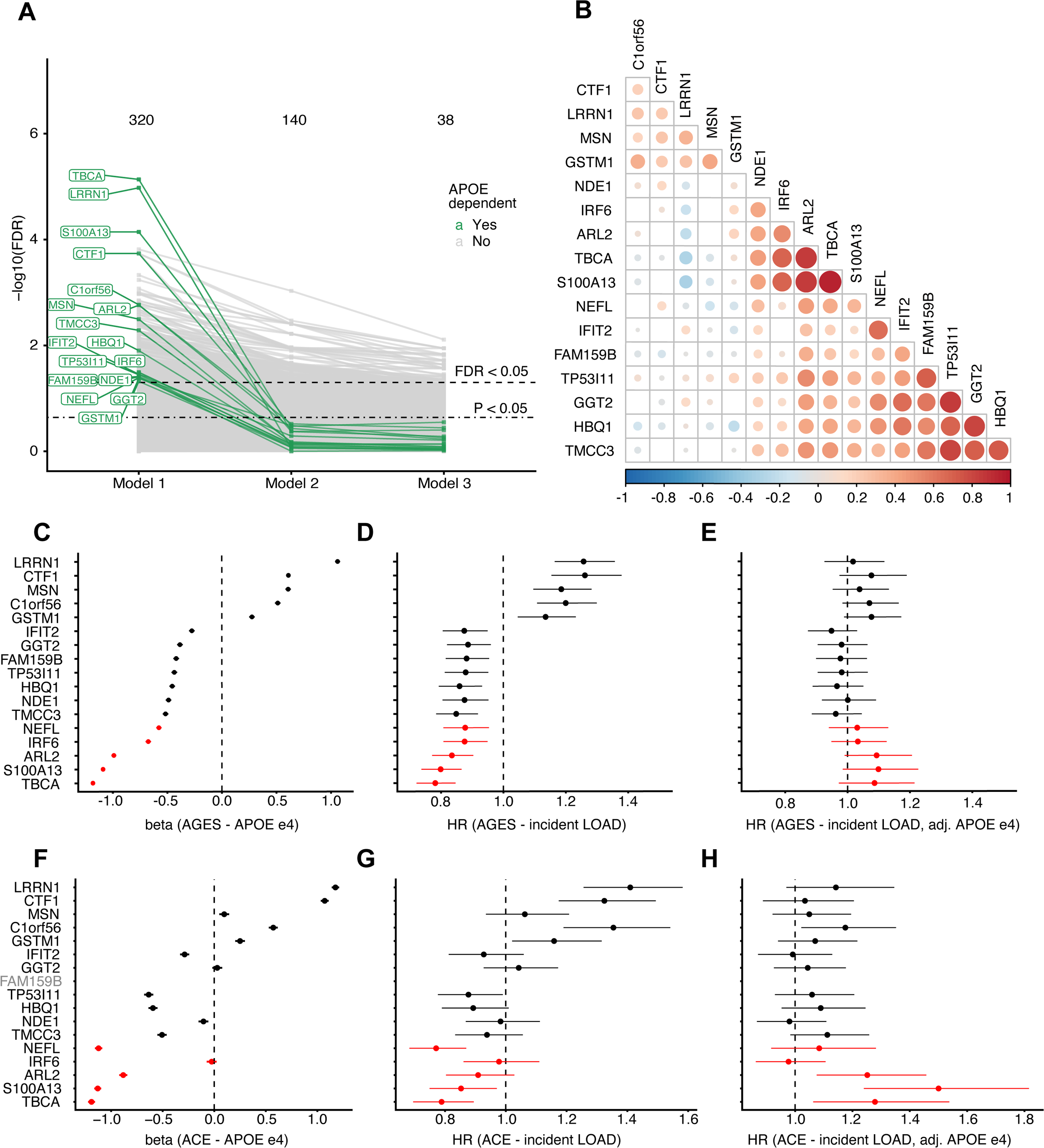
Proteins with *APOE*-ε*4* dependent association to incident LOAD. **A)** Spaghetti plot showing the statistical significance of protein associations with incident LOAD across the three models, highlighting a set of 17 unique proteins (green) whose association with incident LOAD is attenuated upon *APOE-*ε*4* adjustment. The horizontal lines indicate FDR < 0.05 (dashed) and P < 0.05 (dot-dashed). **B)** Pairwise Pearson’s correlation between the 17 *APOE*-dependent proteins. **C)** Forest plot showing the linear associations between *APOE* genotype and the 17 *APOE*-dependent proteins. The beta coefficient shows the change in protein levels per ε4 allele count. **D-E)** Forest plots showing the associations between the 17 *APOE*-dependent proteins and incident LOAD **D)** without *APOE-*ε*4* adjustment (model 1) and **E)** with *APOE-*ε*4* adjustment (model 2). LOAD-HR indicates risk per SD increase of protein levels. Proteins that change direction of effect between the two models are highlighted in red.

The HR conferred by *APOE-*ε*4* for incident LOAD in AGES was 2.1 (P = 1.23e-27) per each ε*4* allele. To evaluate if any of the 17 *APOE*-dependent proteins might mediate the effect of APOE-ε*4* on incident LOAD, we considered the change in HR for *APOE-*ε*4* on risk of incident LOAD when adjusting for individual proteins. We found that adjustment for most proteins resulted in a minor effect decrease, suggesting they do not mediate the *APOE-*ε*4* effect on LOAD (Supplementary Figure 5A). Intriguingly, however, the adjustment for four proteins (NEFL, ARL2, TBCA and S100A13) caused an increase of ∼10% in *APOE-*ε*4* effect size (Supplementary Figure 5A-B). Thus, the effect of *APOE-*ε*4* on LOAD is partly masked by secondary opposing associations between these proteins and LOAD, which are further explored below. The effect of *APOE-*ε*4* carrier status on LOAD risk was largely unchanged in a multivariable model containing all 17 *APOE*-dependent proteins, thus not supporting a mediating effect of these proteins for the LOAD risk conferred by *APOE-*ε*4* (base model: HR = 2.1, P = 1.23e-27, multivariable model: HR = 2.2, P = 3.2e-10). However, although not direct mediators, the 17 proteins could be blood-based readouts of a true mediator within tissue-specific pathological processes occurring prior to LOAD diagnosis.

To map out potential tissues of origin for the circulating levels of the 17 *APOE*-dependent proteins, we considered gene expression data from the Human Protein Atlas^31^. We observed that five (LRRN1, TMCC3, FAM159B, NEFL, GSTM1) of the *APOE*-dependent proteins had elevated gene expression in brain compared to other tissues and additional two (IFIT2, NDE1) clustered with brain-specific genes (Table 2). Of the remaining *APOE*-dependent proteins, six were universally expressed, including in brain tissue, and four were enriched in other tissues. We did not detect any significantly enriched molecular signatures nor GO terms for the 17 *APOE-*dependent proteins (Supplementary Table 7). However, a network analysis of measured and inferred physical protein-protein interactions^32^ revealed that the *APOE*-dependent proteins interact directly with proteins involved in neuronal response- and development, neuroinflammation and AD (Figure 4, Supplementary Table 10-12, Supplementary Note 3).

**Fig. 4.**
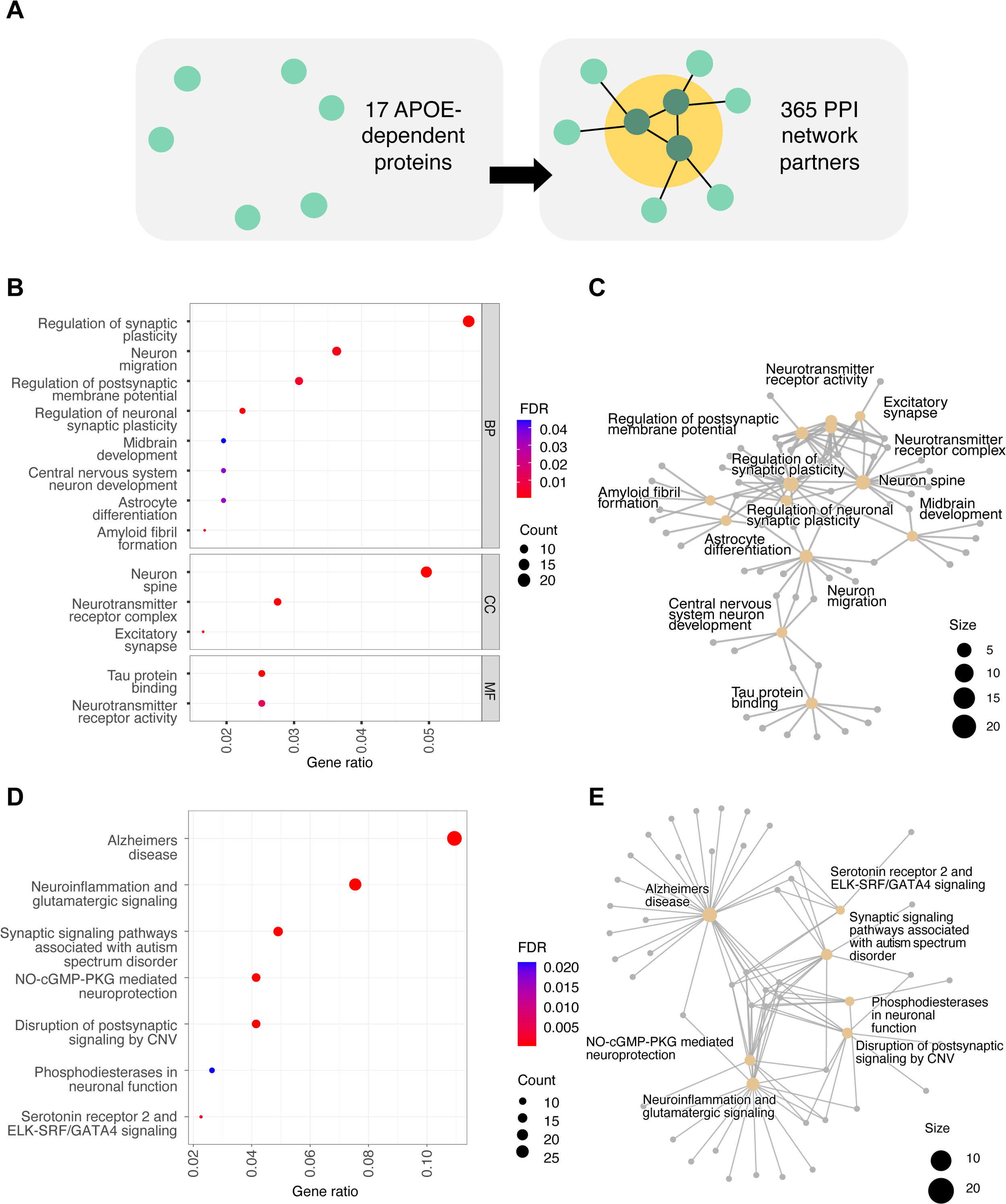
Functional enrichment analysis of *APOE*-dependent protein-protein interaction partners. **A)** A scheme of the PPI partners selection, where first degree partners of the *APOE*-dependent proteins were extracted from the InWeb database. **B-C)** Enrichment of selected Gene Ontology terms for the PPI partner proteins shown as **B)** dotplot and **C)** gene-concept network. **D-E)** Enrichment of top seven unique Wikipathways shown as **D)** dotplot and **E)** gene-concept network.

Given the well-established relationship between APOE and cholesterol^33^ we explored the potential effect of serum lipid levels on the association between LOAD and the 17 *APOE-*dependent proteins (Supplementary Table 13, Supplementary Figures 6-7, Supplementary Note 4). Our findings suggest that, while many of the *APOE*-dependent proteins are associated with cholesterol levels, it is not the driver of their link to LOAD.

### External validation of protein associations with incident LOAD in the ACE cohort

We set to externally evaluate our observations in an independent cohort, the ACE - Alzheimer Center Barcelona (n=1,341), with SOMAscan platform (7k) measurements from plasma of individuals who were referred to the center. The longitudinal component of ACE consists of individuals who had been diagnosed with mild cognitive impairment (MCI) at the center and had been followed up. A total of 719 participants had follow-up information and 266 converted to LOAD over a median follow-up of 3.14 years (Supplementary Table 14). Despite the fundamentally different cohorts, with AGES being population-based and using the 5K SOMAscan platform and ACE being based on individuals with established symptoms and the 7K SOMAscan platform, we replicated 36 protein associations with LOAD at nominal significance (P < 0.05) in the smaller ACE cohort (Table 3, Figure 2F-G). Of those, 30 proteins were nominally significant in model 1 with 97% being directionally consistent with the observations in AGES (Figure 2F). In model 2, 21 proteins were nominally significant, 86% of which were directionally consistent (Figure 2G). After multiple testing correction, seven proteins remained statistically significant (FDR < 0.05), all of which were directionally consistent (Table 3, Figure 2F-G). Six were statistically significant (FDR < 0.05) in model 1 (NEFL, LRRN1, TBCA, CTF1, C1orf56 and TIMP4) and one in model 2 (S100A13) (Supplementary Table 9). Of all 332 tested aptamers, 213 (64%) were directionally consistent regardless of significance in model 1 (Exact binomial test P = 2.0e-05) and 202 (61%) were directionally consistent in model 2 (Exact binomial test P = 0.002), demonstrating an enrichment for consistency in direction of effect. The protein associations replicated in the ACE cohort are of particular interest as they represent potentially clinically relevant candidates for LOAD that are consistent in two different contexts, in both a general population and a clinically derived symptomatic sample set. However, our results suggest that many of the proteins that associate with long-term LOAD risk are not strongly associated with the conversion from MCI to AD, which is further into the AD trajectory and may also explain the limited overlap between the proteins associated with prevalent and incident LOAD in AGES.

**Table 3.**
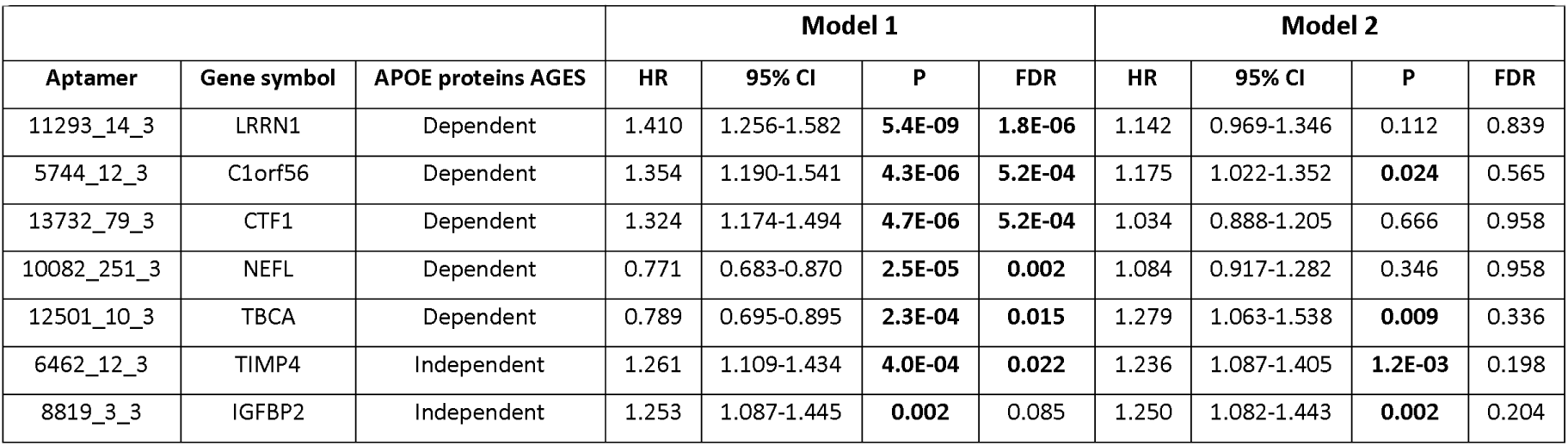
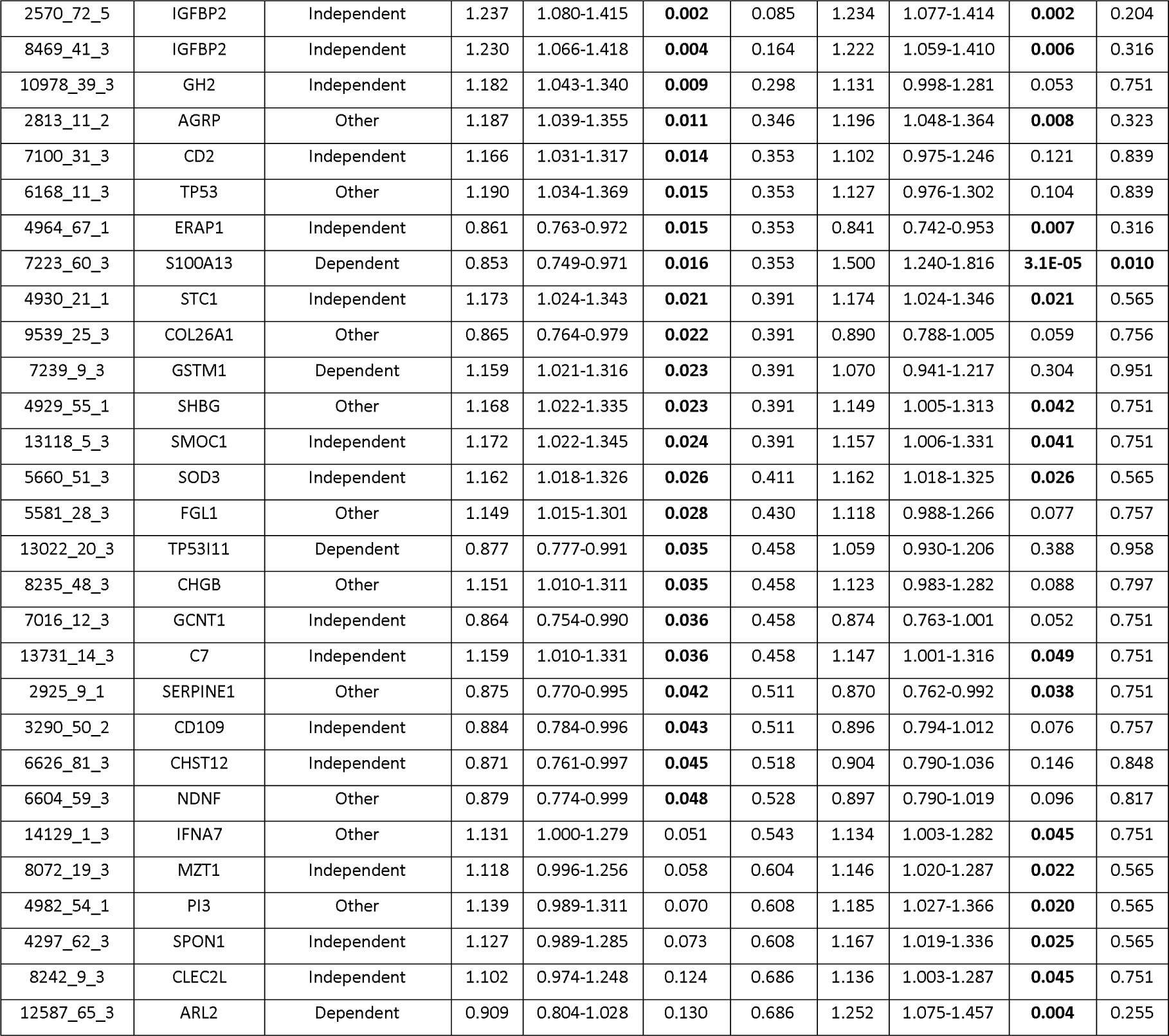
Replication of the LOAD associated proteins from AGES in the ACE cohort. All proteins with nominal P<0.05 in either model are shown. P and FDR values <0.05 are highlighted in bold.

### External validation of reversed LOAD association conditional on APOE-ε4 for a subset of proteins

Specifically considering the *APOE*-dependent proteins, the association between the *APOE-*ε*4* allele and the proteins was replicated for 13 of 17 proteins in the ACE cohort (Figure 3F). Furthermore, the change in direction of effect for incident LOAD upon *APOE-*ε*4* adjustment was replicated in the ACE cohort for 4 of 5 proteins (ARL2, NEFL, S100A13 and TBCA) (Figure 3G-H) (Supplementary Table 9), with even larger effects observed in the ACE cohort compared to AGES in the *APOE-*ε*4* adjusted model and three proteins (ARL2, S100A13 and TBCA) becoming statistically significant (P < 0.05). Thus, the attenuation of the primary LOAD associations for these proteins upon *APOE-*ε*4* adjustment meet the criteria of *APOE-e4* dependence (see Methods). No significant interaction between protein and *APOE-*ε*4* carrier status on AD risk was observed in either the AGES or ACE cohorts. Taken together, our results show that these proteins are strongly downregulated by *APOE-*ε*4,* and consequently show an inverse relationship with incident LOAD, but when adjusting for the *APOE-*ε*4* allele, their association to LOAD is still significant but reversed – suggesting a secondary non-*APOE-*ε*4-*mediated process affecting these same proteins in relation to LOAD in the opposite direction that is more strongly observed a cohort of individuals with MCI than in the population-based AGES cohort.

### Mendelian randomization to identify potential causal associations between proteins and LOAD

The proteins associated with LOAD could include proteins causally related to the disease, or proteins whose serum level changes reflect a response to prodromal or genetic liability to LOAD. To test this hypothesis, we performed a bi-directional two-sample MR analysis, including the targets of all 346 aptamers associated with LOAD in our study. Genetic variant associations for serum protein levels were obtained from a catalog of cis-protein quantitative trait loci (pQTLs) from AGES^24^ while variant associations with LOAD were extracted from a recent GWAS on 39,106 clinically diagnosed LOAD cases, 46,828 proxy-LOAD and dementia cases and 401,577 controls of European ancestry^19^. In total, 117 (34%) of the LOAD-associated serum aptamers had cis-pQTLs that were suitable as genetic instruments and were included in the protein-LOAD MR analysis (Supplementary Table 15).

In the forward MR analysis, two proteins, integrin binding sialoprotein (IBSP) and amyloid precursor protein (APP), had support for causality (Supplementary Table 16). IBSP had a risk-increasing effect for LOAD in both the causal (OR = 1.26, FDR = 0.03) and observational analysis (incident LOAD full follow up, HR = 1.13, FDR = 0.04). APP had a protective effect for LOAD in both the causal (OR = 0.76, FDR = 0.03) and observational analysis (incident LOAD full follow up, HR = 0.87, FDR = 0.047). Notably, while not statistically significant, we observed suggestive support for a protective effect of genetically determined serum levels of acetylcholinesterase (ACHE, OR = 0.92, P = 0.061), a target of clinically used therapeutic agent for dementia^34^ (Supplementary Table 16, Supplementary Figure 8). In a forward MR analysis of the *APOE*-dependent protein interaction partners, two proteins, APP and MAPK3, had support for causality (Supplementary Tables 10-12, Supplementary Note 3).

As most of the observational protein associations in the current study were detected for incident LOAD, and thus reflect changes that take place before the onset of clinically diagnosed disease, it is unlikely that their levels and effects are direct downstream consequences of the disease after it reaches a clinical stage. However, they may reflect a response to a prodromal stage of the disease. We therefore performed a reverse MR to test if the observed changes in serum protein levels are likely to occur downstream of the genetic liability to LOAD, which may capture processes both at the prodromal and clinical stage. The *APOE* locus is likely to have a dominant pleiotropic effect in the reverse MR analysis (Supplementary Table 17, Supplementary Figure 9, Supplementary Note 5), as it has a disproportionately strong effect on LOAD risk compared to all other common genetic variants, while also being a well-established pQTL trans-hotspot, affecting circulating levels of up to hundreds of proteins^24,25,27^. We therefore performed the primary reverse MR analysis using only LOAD-associated genetic variants outside of the *APOE* locus as instruments. We found two proteins (S100A13 and ARL2) that were significantly (FDR < 0.05) affected by LOAD or its genetic liability (Supplementary Table 17, Supplementary Figures 9-10). Interestingly, both were among the 17 previously identified *APOE*-dependent LOAD proteins, together with two additional proteins that were nominally significant in the reverse MR (TBCA, P = 4.4e-4, FDR = 0.051 and IRF6, P = 7.9e-4, FDR = 0.055). Thus intriguingly, these findings suggest that these four proteins are upregulated by LOAD, in contrast to the observed *APOE-*ε*4* downregulation of the same proteins (Figure 5). This supports our findings of competing biological effects described above (Figure 3E, Supplementary Figure 5) and collectively our results indicate that simultaneous opposing effects of *APOE-*ε*4* on one hand and LOAD on the other result in differential regulation of these proteins in serum (Figure 5B).

**Fig. 5.**
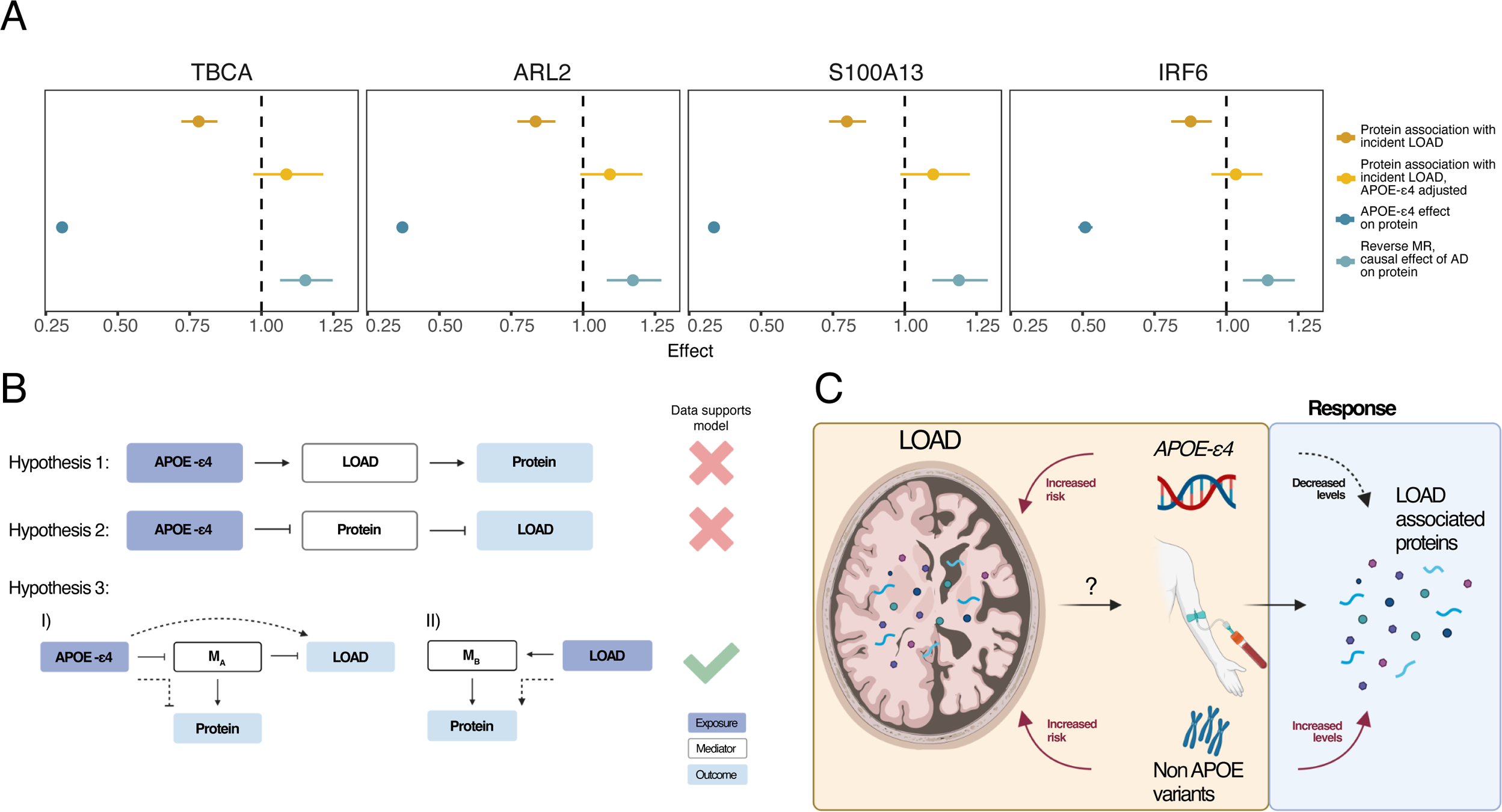
Reverse Mendelian randomization analysis. **A)** Comparison of hazard ratios for incident LOAD with and without *APOE-*ε*4* adjustment in the observational analysis, the effects of *APOE-*ε*4* on protein levels and reverse MR odds ratios (excluding the *APOE* locus) for the four *APOE*-dependent proteins that change direction of effect in both observational and causal analyses when *APOE* is accounted for. **B-C)** Visual summaries of the observed data. **B)** Mediation diagrams showing 3 possible hypotheses that could explain the relationship between *APOE-*ε*4,* LOAD and the four proteins shown in **A)**. Our analyses do not support the hypothesis that LOAD mediates the effect of *APOE-*ε*4* on proteins (Hypothesis 1) nor the other way around (Hypothesis 2). However, our results from both the observational and causal analyses support the hypothesis that two mechanisms are at play that affect the same proteins in the opposite direction (Hypothesis 3). **C)** The *APOE-*ε*4* mutation leads to increased risk of LOAD via its effects in brain tissue. The same mutation results in a downregulation of serum levels of four proteins that are themselves negatively associated with incident LOAD. Additionally, other non-*APOE* LOAD risk variants lead to upregulation of the same proteins in the reverse MR analysis, possibly reflecting a response to LOAD or its genetic liability.

We performed a replication analysis of the effect of *APOE-*ε*4* on protein levels and the reverse MR results for these four proteins using published protein GWAS summary statistics from two recent studies^25,35^. In the external datasets, the downregulation of all four proteins by *APOE-*ε*4* (as determined by the rs429358 C allele) was replicated. In the reverse MR analysis (excluding the *APOE* locus), the upregulation of protein levels by LOAD liability observed in AGES was also detected for two proteins (S100A13 and TBCA) in both validation cohorts, reaching significance (P < 0.05) in the study by Ferkingstad et al. (Supplementary Figure 11, Supplementary Table 18). While the two proteins changed direction in a similar manner as in AGES, the effect size was considerably smaller in the validation cohorts. Importantly, however, individuals in these two cohorts are much younger than those in AGES, with mean ages of 55 and 48 years for the Ferkingstad et al. and Sun et al. studies, respectively, compared to 76 years in AGES. Therefore, we conducted an age-stratified reverse MR analysis in AGES, which showed a strong age-dependent effect, with a much larger effect of LOAD genetic liability on protein levels in individuals over 80 years old compared to those younger than 80 years (Supplementary Figure 11). The effect size in AGES individuals under 80 years old was in line with the effect observed in the validation cohorts. Thus, if the upregulation of these proteins reflects a response to prodromal or preclinical LOAD, an older cohort may be needed to detect an association of the same degree as we found in AGES. However, the observed support in the validation cohorts for the discordant effects of *APOE* vs non-*APOE* LOAD-associated genetic variants on the same serum proteins strongly implicates these proteins as directly relevant to LOAD, potentially as readouts of biological processes that are both disrupted by *APOE-*ε*4* and modulated in the opposite manner as a response to genetic predisposition to LOAD or the disease onset in general.

Together, these results indicate that LOAD or its general genetic liability causally affects the levels of some *APOE*-dependent proteins, but this effect is simultaneously masked by the strong effects of the *APOE* locus in the other direction (Figure 5A). These outcomes strengthen results described above, showing that the levels of these four proteins are strongly downregulated in *APOE-*ε*4* carriers and lower levels of these proteins are therefore associated with increased risk of LOAD in an *APOE*-dependent manner (Figure 5B). Simultaneously, the reverse MR analysis shows that the collective effect of the other non-*APOE* LOAD risk variants is to upregulate the serum levels of these same proteins, possibly reflecting a response mechanism to LOAD pathogenesis (Figure 5C). Again, this is in line with the observational analysis, where all four proteins changed direction of effect when adjusting for *APOE-*ε*4* (Figure 5A, Figure 2D-E).

### Overlap with the AD brain and CSF proteome

To evaluate to what extent our LOAD-associated serum proteins reflect the proteomic profile of AD in relevant tissues, we queried data from recent proteomic studies of AD in cerebrospinal fluid (CSF)^36^ and brain^37^ which also describe tissue specific co-regulatory modules. We observed that of our LOAD-associated serum proteins, 51 proteins were also associated with AD in brain as measured by mass-spectrometry, with 32 (63%) being directionally consistent (Figure 6A-B) (Supplementary Tables 19-20). Higher directional consistency was observed within the *APOE*-independent protein group, or 15 (71%) of 21 proteins associated with AD in brain tissue. Additionally, 60 proteins were directly associated with AD in CSF as measured with SOMAscan (7k) (Figure 6A) with 46 (77%) being directionally consistent (Figure 6B)^21^. The proportion of directionally consistent associations between serum and CSF was higher in both the *APOE*-independent and dependent protein groups, or 88% (22 of 25 and 7 of 8 for *APOE*-independent and dependent proteins, respectively) (Figure 6B, Supplementary Table 19). However, directional inconsistency between plasma and CSF AD proteomic profiles has been reported before in a similar comparison^38^. Fourteen proteins overlapped between all three tissues in the context of AD (Figure 6A) (Supplementary Table 19). Many of these proteins have established links or are highly relevant to LOAD, such as Spondin 1 (SPON1), involved in the processing of amyloid precursor protein (APP)^39^; Secreted Modular Calcium-Binding Protein 1 (SMOC1) previously proposed as a biomarker of LOAD in postmortem brains and CSF^40^; Netrin-1 (NTN1), an interactor of APP and regulator of amyloid-beta production^41^; Neurofilament light (NEFL), previously proposed as a plasma biomarker for LOAD and axon injury^42,43^ and Von Willebrand factor (VWF), known for its role in blood clotting and associations with LOAD^44^ (Supplementary Table 19). Notably, some of the *APOE*-dependent proteins were associated with AD across all three tissues such as TBCA and TP53I11.

**Fig. 6.**
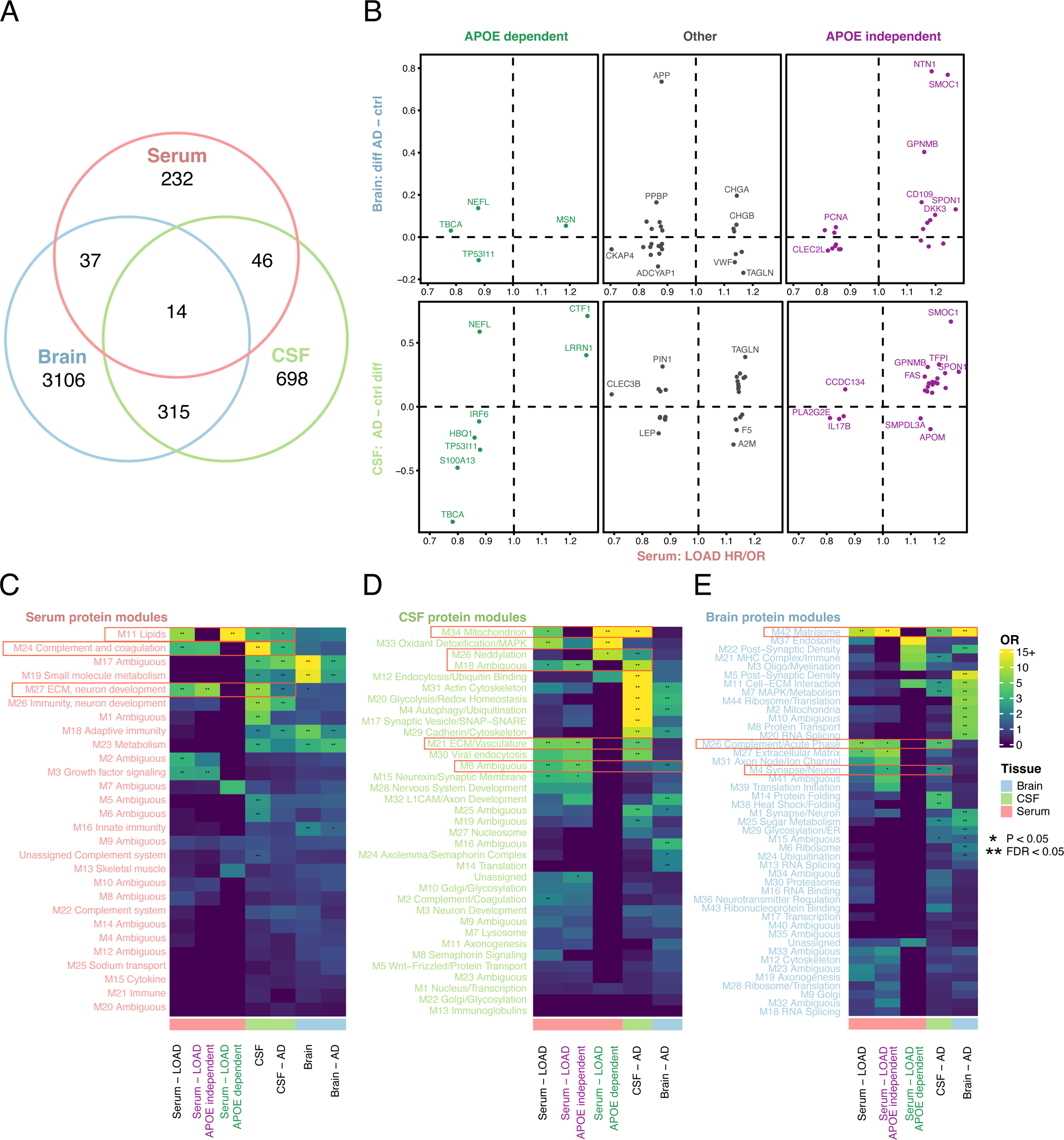
Overlap between AD protein signatures in serum, brain and CSF. **A)** A Venn diagram showing the overlap of AD-associated proteins in serum, brain and CSF. **B)** A comparison of the effect sizes for AD associated proteins that overlap between serum and brain (top) and serum and CSF (bottom). The proteins are stratified based on the APOE-dependence in AGES for incident LOAD. The effect size in AGES is shown for incident LOAD model 1, except for proteins that were uniquely identified using the shorter 10-year follow-up or prevalent LOAD, in which case the respective effect size from the significant association is shown. **C-E)** Heatmap showing the enrichment of AD-associated proteins by tissue type (x-axis) in **C)** the AGES serum protein modules, **D)** Emory CSF protein modules and **E)** Emory brain protein modules (y-axis).

We have previously described the co-regulatory structure of the serum proteome, which can broadly be defined as 27 modules of correlated proteins^26^ (Supplementary Table 21). In the current study we found that among the 346 aptamers (329 proteins) associated with LOAD (prevalent or incident, any model), five serum protein modules (M27, M3, M11, M2 and M24) were overrepresented (Figure 6C, Supplementary Table 22). In particular, the 140 *APOE*-independent aptamers were specifically overrepresented in module M27, enriched for proteins involved in neuron development and the extracellular matrix, and module M3 that is associated with growth factor signaling pathways (Supplementary Table 22). By contrast, the 17 *APOE-*dependent proteins were specifically enriched in protein module M11 (Supplementary Table 22), which is strongly enriched for lipid pathways and is under strong genetic control of the *APOE* locus^26^. Serum modules M27, M24 and M11 were all enriched for AD-associations in CSF (Figure 6C). We next sought to understand to what extent our LOAD-associated proteins identified in serum might reflect AD protein signatures in CSF and brain tissue. Among the LOAD-associated proteins measured in serum, we found the *APOE*-dependent and *APOE*-independent proteins to be enriched in different CSF modules, most of which were also linked to AD (Figure 6D, Supplementary Table 22). In brain tissue, the serum *APOE*-independent LOAD proteins were particularly enriched in brain module M42 (Matrisome), which is enriched for extracellular matrix (ECM) proteins^37^. Strikingly, M42 was strongly enriched for the AD-proteomic profiles of all three tissues (Figure 6E, Supplementary Table 22). Interestingly, members of this module (SMOC1, APP, SPON1, NTN1, GPNMB) showed some of the strongest associations in serum to incident LOAD in our study (Figure 2D, Supplementary Table 2) as well as in brain (Figure 6B, Supplementary Table 22). This module has furthermore been demonstrated to be correlated with amyloid beta (Aβ) deposition in the brain and some of its protein constituents (e.g MDK, NTN1 and SMOC1) have been shown to colocalize with and bind to Aβ^37^. Additionally, the *APOE* locus regulates M42 levels in the brain (mod-qTL), and while the APOE protein is a member of module M42, this regulation was found to not be solely driven through the levels of the APOE protein itself^37^. Our results simultaneously show that other members of the module, such as SPON1 and SMOC1, exhibit an *APOE*-independent association to incident LOAD in serum. Interestingly, these same two proteins are increased in CSF thirty years prior to symptom onset in autosomal dominant early onset AD^45^. In summary, we demonstrate significant overlaps in LOAD-associated protein expression across blood, CSF and brain on both an individual protein level and on protein module level.

## Discussion

We describe a comprehensive mapping of the serum protein profile of LOAD that provides insight into processes that are independent or dependent on the genetic control of *APOE*-ε*4* (Figure 7). We identified 329 proteins in total that differed in the incident or prevalent LOAD cases compared to non-LOAD participants in a population-based cohort with long-term follow-up. Among these, we identified a novel grouping of proteins based on their primary LOAD-association being statistically independent of (130 proteins), or dependent on (17 proteins) *APOE*-ε*4* carrier status. Many of the *APOE*-independent proteins are implicated in neuronal pathways and are shared with the LOAD-associated CSF and brain proteome. The 17 *APOE*-dependent proteins overlap with AD-associated protein modules in CSF and interact directly with protein partners involved in LOAD, including APP. Another key finding is, amongst these 17 proteins, four proteins change LOAD-associated direction of effect both observationally and genetically when taking *APOE-*ε*4* carrier status into account. Collectively, our results suggest that while their primary association with LOAD reflects the risk conferred by *APOE*-ε*4*, there exists a secondary causal effect of LOAD itself on the protein levels in the reverse direction as supported by the MR analysis, possibly reflecting a response to the disease onset.

**Fig. 7.**
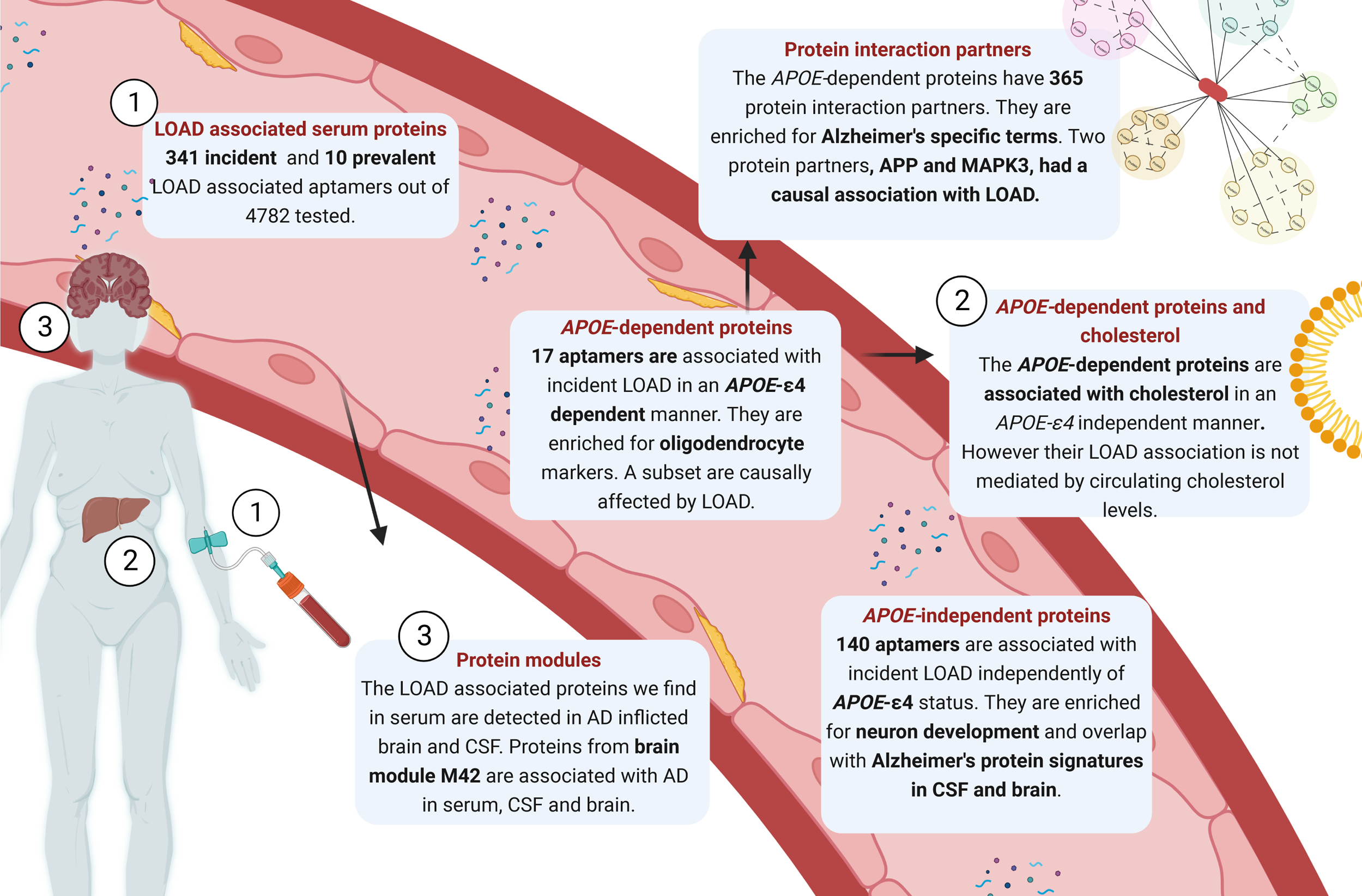
Graphical summary of the results.

Previous studies identifying proteins associated with LOAD have been limited to a cross-sectional cohort or are based on all-cause dementia^18,46–48^. Here we extend those findings by distinguishing LOAD cases from other types of dementia in a prospective cohort study to identify LOAD-specific serum protein signatures preceding clinical onset. Furthermore, our comparative approach of statistical models with and without *APOE-*ε*4* adjustment provides a novel compartmentalized view of the LOAD serum protein profile and demonstrates how protein effects can differ depending on genetic confounders which are imperative to take into consideration. We found that the proteins associated with incident LOAD in our study, in particular those independently of *APOE-*ε*4* such as GPNMB, NTN1, SMOC1 and SPON1, overlap with the proteomic profile of LOAD in CSF^38^ and brain^37^, and are enriched for neuronal pathways, which may reflect an altered abundance of neuronal proteins in the circulation during the prodromal stage of LOAD. These overlaps that we find across independent cohorts and different proteomics technologies suggest that the serum levels of some proteins have a direct link to the biological systems involved in LOAD pathogenesis and may even provide a peripheral readout of neurodegenerative processes prior to clinical diagnosis of LOAD. In particular, the proteins that show directionally consistent effect sizes suggest exceptional AD-specific robustness as the measurements vary by tissue, methodology and populations.

We identified 17 proteins with a particularly strong *APOE*-dependent association to incident LOAD, of which eight were also associated with prevalent AD in CSF. The association between *APOE-*ε*4* and circulating levels of these proteins has been reported by our group^24,26,27^ and others^49^, but their direct association with incident LOAD has to our knowledge not been previously described. These *APOE*-dependent proteins may point directly to the processes through which *APOE-*ε*4* mediates its risk on LOAD and provide a readout of the pathogenic process in the circulation of the approximately 50% of LOAD patients worldwide carrying the variant^21,22^. While our data does not provide information on the tissue-origin of the *APOE*-dependent proteins, nine either exhibit brain-specific gene expression, cluster with brain-specific genes^50^ or have been associated with LOAD at the transcriptomic or protein level in brain tissue or CSF (Table 2). At the genetic level, a lookup in the GWAS catalog^51^ shows that an intron variant in the *IRF6* gene has a suggestive GWAS association with LOAD via *APOE-*ε*4* carrier status interaction^52^. In addition, variants in the *TMCC3* gene have been linked to educational attainment^53^ and caudate volume change rate^54^ and variants in the *TBCA* gene have been suggestively associated with reaction time^55^ and PHF-tau levels^56^. Collectively, the gene expression patterns for these proteins in the brain, interactions with proteins involved in neuronal processes and suggestive associations between genetic markers in or near these genes and brain-related outcomes suggest that these *APOE-*dependent proteins may reflect brain-specific processes affected by *APOE-*ε*4* carrier status that affect the risk of developing LOAD. Importantly, the association patterns for ARL2, S100A13 and TBCA suggest the presence of a pathway that is downregulated by *APOE-*ε*4* early in life, given the consistent effect of *APOE-*ε*4* on the same proteins in younger cohorts, but upregulated at the onset of LOAD, as supported by the larger observed effects in the *APOE-*ε*4* adjusted analysis in the ACE cohort of individuals who are closer to diagnosis on the AD trajectory than those in AGES. Additional studies are required to expand on these interpretations and dissect the complex mechanisms at play and to determine if the modulation of the process represented by these proteins has therapeutic potential.

Two proteins, IBSP and APP, were identified to potentially have a causal role in LOAD. IBSP was previously associated with plasma amyloid-β and incident dementia^57^, while APP is the precursor protein for amyloid-β^58^. Based on the MR analysis for the LOAD-associated proteins that could be tested, the majority do not appear to be causal in and of themselves but their association with incident LOAD may still reflect changes that occur years before the onset of LOAD that could be of interest to target before irreversible damage accumulates.

A major strength of this study is the high-quality data from a prospective longitudinal population-based cohort study with detailed follow-up, broad coverage of circulating proteins and a comprehensive comparison to the AD-proteome in CSF and brain. The limitations of our study include that our results are based on a Northern European cohort and cannot necessarily be transferred directly to other populations or ethnicities. Additionally, while we partly replicate our overall findings in an external cohort, a greater replication proportion could be anticipated in a more comparable cohort. The ACE cohort consists of clinically referred individuals with MCI and proteomic measurements performed on a different version of the SOMAscan platform. Additionally, different normalization procedures were applied by SomaLogic for the two SOMAscan versions, which may have an effect on the LOAD associations^48^. Further studies are required to determine the impact of time to event, platform and normalization approaches on the associations between circulating proteins and LOAD. Regardless of these differences, we did replicate the majority of the *APOE*-dependent LOAD associations, including the *APOE*-dependent change in effect for ARL2, S100A13 and TBCA. We could not test all LOAD-associated proteins for causality, including most of the *APOE*-dependent proteins, due to lack of significant cis-pQTLs for two thirds of the proteins, thus we cannot exclude the possibility that some could be causal but missed by our analysis. Finally, despite our LOAD diagnosis criteria it is possible that some of our findings reflect processes related to dementia in general. As a result, it is critical that these findings be validated in individuals with established amyloid-beta and tau deposits, as well as in experimental settings.

The proteins highlighted in this study and the mechanisms they point to may be used as a source of biomarkers or therapeutic targets that can be modulated for the prevention or treatment of LOAD. This large prospective cohort study, using both a longitudinal and cross-sectional design, represents a unified and comprehensive reference analysis with which past and future serum protein biomarkers and drug targets can be considered, compared, and evaluated.

## Supporting information

Supplementary meterial

## Data Availability

All data produced in the present study are available upon reasonable request to the authors

## Acknowledgements

The authors acknowledge the contribution of the Icelandic Heart Association (IHA) staff to AGES-Reykjavik, as well as the involvement of all study participants. The protein measurements in AGES were supported by Novartis Biomedical Research. The National Institute on Aging (NIA) contracts N01-AG-12100 and HHSN271201200022C for V.G. financed the study. IHA received a grant from Althingi (the Icelandic Parliament), V.G. received funding from the NIA (1R01AG065596-01A1) and E.J. from the NIA (1K08AG068604). A.I.L was also funded from the NIA (P30AG066511 and U01AG061357). The content is solely the responsibility of the authors and does not necessarily represent the official views of the National Institutes of Health. The Genome Research @ Ace Alzheimer Center Barcelona project (GR@ACE) is supported by Grifols SA, Fundación bancaria ‘La Caixa’, Ace Alzheimer Center Barcelona and CIBERNED. Ace Alzheimer Center Barcelona is one of the participating centers of the Dementia Genetics Spanish Consortium (DEGESCO). The FACEHBI study is supported by funds from Ace Alzheimer Center Barcelona, Grifols, Life Molecular Imaging, Araclon Biotech, Alkahest, Laboratorio de análisis Echevarne and IrsiCaixa. Authors acknowledge the support of the Spanish Ministry of Science and Innovation, Proyectos de Generación de Conocimiento grants PID2021-122473OA-I00, PID2021-123462OB-I00 and PID2019-106625RB-I00. ISCIII, Acción Estratégica en Salud, integrated in the Spanish National R+D+I Plan and financed by ISCIII Subdirección General de Evaluación and the Fondo Europeo de Desarrollo Regional (FEDER “Una manera de hacer Europa”) grants PI13/02434, PI16/01861, PI17/01474, PI19/00335, PI19/01240, PI19/01301, PI22/01403, PI22/00258 and the ISCIII national grant PMP22/00022, funded by the European Union (NextGenerationEU). The support of CIBERNED (ISCIII) under the grants CB06/05/2004 and CB18/05/00010. The support from the ADAPTED and MOPEAD projects, European Union/EFPIA Innovative Medicines Initiative Joint (grant numbers 115975 and 115985, respectively); from PREADAPT project, Joint Program for Neurodegenerative Diseases (JPND) grant N° AC19/00097; from HARPONE project, Agency for Innovation and Entrepreneurship (VLAIO) grant N° PR067/21 and Janssen. DESCARTES project is funded by German Research Foundation (DFG). Amanda Cano received support from the ISCIII under the grant Sara Borrell (CD22/00125).

## Author contributions

Conceptualization – E.A.F., Va.G., V.E, Vi.G.

Formal Analysis – E.A.F., Va.G., T.J., A.E.S., E.F.G., A.G., T.A., E.B.D., A.S.

Resources – Vi.G., V.E., A.I.L., A.R., A.P.O., J.J.L., L.L.J., N.T.S., E.C.B.J., L.J.L.

Data Curation – R.P., A.C., M.B., P.G.G., S.V., Va.G., T.A., E.F.G.

Writing original draft – E.A.F., Va.G.

Writing review & editing – All authors

Visualization – E.A.F., Va.G.

Supervision – Va.G., Vi.G.

Funding Acquisition – Vi.G., L.J.L.

## Methods

### AGES study population

Participants aged 66 through 96 were from the Age, Gene/Environment Susceptibility (AGES)-Reykjavik Study cohort. AGES is a single-center prospective population-based study of deeply phenotyped subjects (n = 5,764, mean age 76.6⍰±⍰5.6 years) and survivors of the 40-year-long prospective Reykjavik study, an epidemiologic study aimed to understand aging in the context of gene/environment interaction by focusing on four biologic systems: vascular, neurocognitive (including sensory), musculoskeletal, and body composition/metabolism^30^. Of the AGES participants, 3,411 attended a 5-year follow-up visit. LOAD diagnosis at AGES baseline and follow-up visits was carried out using a three-step procedure described in Sigurdsson et al.^67^. Cognitive assessment was carried out on all participants. Neuropsychological testing was performed on individuals with suspected dementia. Individuals remaining suspect for dementia underwent further neurologic and proxy examinations in the second diagnosis step. Thirdly, a panel comprising of a neurologist, geriatrician, neuroradiologist, and neuropsychologist assessed the positive scoring participants according to international guidelines^30^ and gave a dementia diagnosis. The participants were followed up for incident dementia through medical and nursing home reports and death certificates. The follow-up time was up to 16.9 years, with the last individual being diagnosed 16 years from baseline. Nursing home reports were based on intake exams upon entry or standardized procedures carried out in all Icelandic nursing homes^68^. The participants diagnosed at baseline were defined as prevalent LOAD cases while individuals diagnosed with LOAD during the follow-up period were defined as incident LOAD cases. All prevalent non-AD dementia cases (n=163) were excluded from analyses.

Age, sex, education, and lifestyle variables were assessed via questionnaires at baseline. Education was categorized as primary, secondary, college, or university degree. Smoking was characterized as current, former, or never smoker. *APOE* genotyping was assessed via microplate array diagonal gel electrophoresis (MADGE)^69^. BMI and hypertension were assessed at baseline. BMI was calculated as weight (kg) divided by height (m) squared, and hypertension was defined as antihypertensive treatment or BP >140/90 mm Hg. Type 2 diabetes was defined from self-reported diabetes, diabetes medication use, or fasting plasma glucose ≥7 mmol/L. Serum creatinine was measured via the Roche Hitachi 912 instrument and estimated glomerular filtration rate (eGFR) derived with the four-variable MDRD study equation^70^. The AGES study was approved by the NBC in Iceland (approval number VSN-00-063), and by the National Institute on Aging Intramural Institutional Review Board, and the Data Protection Authority in Iceland.

### Proteomic measurements

The proteomic measurements in AGES have been described in detail elsewhere^27,71^ and was available for 5,457 participants. Briefly, a custom version of the SOMAscan platform (Novartis V3-5K) was applied based on the slow-off rate modified aptamer (SOMAmer) protein profiling technology^72,73^ including 4,782 aptamers that bind to 4,137 human proteins. Serum was prepared using a standardized protocol^74^ from blood samples were collected after an overnight fast, stored in 0.5 ml aliquots at −80°C and serum samples that had not been previously thawed were used for the protein measurements. All samples were run as a single set at SomaLogic Inc. (Boulder, CO, US). Hybridization controls were used to adjust for systematic variability in detection and calibrator samples of three dilution sets (40%, 1%, and 0.005%) were included so that the degree of fluorescence was a quantitative reflection of protein concentration. All aptamers that passed quality control had median intra-assay and inter-assay coefficient of variation (CV)1<15%. Finally, intraplate median signal normalization was applied to individual samples by SomaLogic instead of normalization to an external reference of healthy individuals, as is done for later versions of the SOMAscan platform (https://somalogic.com/wp-content/uploads/2022/07/SL00000048_Rev-3_2022-01_-Data-Standardization-and-File-Specification-Technical-Note-v2.pdf).

### ACE cohort

ACE Alzheimer Center Barcelona was founded in 1995 and has collected and analyzed roughly 18,000 genetic samples, diagnosed over 8,000 patients, and participated in nearly 150 clinical trials to date. For more details, visit www.fundacioace.com/en. The syndromic diagnosis of all subjects of the ACE cohort was established by a multidisciplinary group of neurologists, neuropsychologists, and social workers. Healthy controls (HCs), including individuals with subjective cognitive decline (SCD) diagnosis, were assigned a Clinical Dementia Rating (CDR) of 0, and mild cognitive impairment (MCI) individuals a CDR of 0.5. For MCI diagnoses, the classification of López *et al.,* 2003, and Petersen’s criteria were used^75–78^. The 2011 National Institute on Aging and Alzheimer’s Association (NIA-AA) guidelines were used for AD diagnosis^79^. All ACE clinical protocols have been previously published^80–82^. Paired plasma and CSF samples^83^, following consensus recommendations, were stored at −80°C. A subset of ACE cohort was analyzed with the SOMAscan 7k proteomic platform^84^ (n = 1,370), (SomaLogic Inc., Boulder, CO, US). The proteomic data underwent standard quality control procedures at SomaLogic and was median normalized to reference using the Normalization by Maximum Likelihood (ANML) method (https://somalogic.com/wp-content/uploads/2022/07/SL00000048_Rev-3_2022-01_-Data-Standardization-and-File-Specification-Technical-Note-v2.pdf). Additionally, *APOE* genotyping was assessed using TaqMan genotyping assays for rs429358 and rs7412 SNPs (Thermo Fisher). Genotypes were furthermore extracted from the Axiom 815K Spanish Biobank Array (Thermo Fisher) performed by the Spanish National Center for Genotyping (CeGen, Santiago de Compostela, Spain).

### Statistical analysis

<u>Protein measurement</u> data was centered, scaled and Box-Cox transformed, and extreme outliers excluded as previously described^71^. Sample size was not predetermined by any statistical method but rather by available data. The associations of serum protein profiles with prevalent AD (n = 167) were examined cross-sectionally via logistic regression at baseline. The associations of serum protein profiles with incident LOAD (n = 655) were examined longitudinally via Cox proportional-hazards models. Participants who died or were diagnosed with incident non-AD dementia were censored at date of death or diagnosis. To account for hazard ratio variability which may arise with lengthy follow-ups, a secondary analysis using 10-year follow-up cut-off of incident LOAD was performed (n_LOAD_ = 432). To compare the fits of the two follow-up times and test for time-dependence of the coefficients we used anova and the survsplit function from the Survival R package^85^. For both prevalent and incident LOAD, we examined three covariate-adjusted models. The primary model (model 1) included the covariates sex and age. Model 2 included as an additional covariate the *APOE-*ε*4* allele count. The third model (model 3) included additional adjustment for cardiovascular risk factors, lifestyle, and kidney function (BMI, type 2 diabetes, education, hypertension, smoking history, eGFR) as they have been associated with risk of LOAD^86^. Benjamini-Hochberg false discovery rate (FDR) was used to account for multiple hypothesis testing. Analyses were conducted using R version 4.2.1. ACE SomaLogic proteomics data was similarly Box-Cox transformed and association analysis performed in the same manner as in AGES.

*<u>APOE</u>*<u>-dependence criteria</u> of the proteins were defined as serum proteins that met FDR significance of < 0.05 in association with incident LOAD in model 1, thus unadjusted for the *APOE-*ε*4* allele, but whose nominal significance was abolished upon *APOE-*ε*4* correction in model 2 (P > 0.05). Serum proteins that remained nominally significantly associated with incident LOAD (P < 0.05) upon *APOE-*ε*4* correction but changed direction of effect were also considered to meet the *APOE* dependence criteria, as a reversal of the effect indicates that the primary association is driven by *APOE-*ε*4*.

<u>Functional enrichment</u> analyses were performed using Over-Representation Analysis (ORA) and Gene Set Enrichment Analysis (GSEA) using the R packages ClusterProfiler and fgsea^87,88^. The association significance cut-off for inclusion in ORA was FDR < 0.05. Background for both methods was specified as all proteins tested from the analysis leading up to enrichment testing. The investigated gene sets were the following: Gene Ontology, Human Phenotype Ontology, KEGG, Wikipathways, Reactome, Pathway Interaction Database (PID), MicroRNA targets (MIRDB and Legacy), Transcription factor targets (GTRD and Legacy), ImmuneSigDB and the Vaccine response gene set^89^. Finally, we looked into tissue gene expression signatures via the same methods (ORA and GSEA) using data from GTEX^90^ and The Human Protein Atlas, where gene expression patterns across tissues were categorized in a similar manner as described by Uhlen et al^50^ and tissue-elevated expression considered as gene expression in any of the categories ‘tissue-specific’, ‘tissue-enriched’ or ‘group-enriched’. MinGSSize was set at 2 when investigating the LOAD-associated serum proteins directly. When investigating the *APOE-*dependent protein interaction partners, minGSSize was set to 15 and maxGSSize was set to 500. Overrepresentation of brain cell type markers among LOAD-associated proteins was tested using a Fisher’s exact test, with the SOMApanel protein set as background. Tissue specificity lookup for the top LOAD associated proteins was based on the Human Protein Atlas version 22 (https://v22.proteinatlas.org/). For the protein-protein interaction (PPI) network analysis, PPIs from InWeb^32^ (n = 14,448, after Entrez ID filtering) were used to obtain the first-degree interaction partners of the *APOE-*dependent proteins.

### Protein comparisons across serum, CSF and brain

To compare protein modules and AD associations across tissues, protein modules and protein associations to AD were obtained from brain^37^ and CSF^36^. The brain data, from the Banner Sun Health Research Institute^91^ and ROSMAP^92^, included TMT-MS-based quantitative proteomics for 106 controls, 200 asymptomatic AD cases and 182 AD cases. The CSF samples were collected under the auspices of the Emory Goizueta Alzheimer’s Disease Research Center (ADRC) and Emory Healthy Brain Study (EHBS)^36^. The cohort consisted of 140 healthy controls and 160 patients with AD as defined by the NIA research framework^93^. Protein measurements were performed using TMT-MS and SomaScan (7k). Only SomaLogic protein measurements were included in the comparison between CSF and serum, which were median normalized. Proteins were matched on SomaLogic aptamer ID when possible but otherwise by Entrez gene symbol. Overlaps between modules and AD-associated (FDR<0.05) proteins across tissues were evaluated with Fisher’s exact test.

### Mendelian randomization

A two-sample bi-directional Mendelian randomization (MR) analysis was performed to first evaluate the potential causal effects of serum protein levels on AD (forward MR), and second to evaluate the potential causal effects of AD or its genetic liability on serum protein levels (reverse MR). All aptamers significantly (FDR<0.05) associated with LOAD (incident or prevalent) were included in the MR analyses, or a total of 346 unique aptamers (Supplementary Tables 2, 3 and 5), of which 320 aptamers were significant in the full follow-up incident LOAD analysis (models 1-3), 106 aptamers were significant in the 10-year follow-up incident LOAD analysis (model 1-3) and ten aptamers were significant in the prevalent LOAD analysis (models 1-3). Genetic instruments for serum protein levels were obtained from a GWAS of serum protein levels in AGES^24^ and defined as follows. All variants within a 11Mb (±5001kb) cis-window for the protein-encoding gene were obtained for a given aptamer. A cis-window-wide significance level Pb1=10.05/N, where N equals the number of SNPs within a given cis-window, was computed and variants within the cis window for each aptamer were clumped (r^2^1≥10.2, P1≥1Pb). The effect of the genetic instruments for serum protein levels on LOAD risk was obtained from a GWAS on GWAS on 39,106 clinically diagnosed LOAD cases, 46,828 proxy-LOAD and dementia cases and 401,577 controls of European ancestry^19^. Genetic instruments for the serum protein levels not found in the LOAD GWAS data set were replaced by proxy SNPs (r^2^1>10.8) when possible, to maximize SNP coverage. Genetic instruments for LOAD in the reverse causation MR analysis were obtained from the same LOAD GWAS^24^, where genome-wide significant variants were extracted (P1<15e-8) and clumped at a more stringent LD threshold (r21≥10.01) than for the protein instruments to limit overrepresentation of SNPs from any given locus across the genome. In the reverse causation MR analysis, cis variants (±500 kb) for the given protein were excluded from the analysis to avoid including pleiotropic instruments affecting the outcome (protein levels) through other mechanisms than the exposure (LOAD). A secondary reverse causation MR analysis was performed excluding any variants in the *APOE* locus (chr19:45,048,858-45,733,201). Causal estimate for each protein was obtained by the generalized weighted least squares (GWLS) method^94^, which accounts for correlation between instruments. Causality for proteins with single cis-acting variants was assessed with the Wald ratio estimator. For the reverse causation MR analysis, the inverse variance weighted method was applied due to a more stringent LD filtering of the instruments. Instrument heterogeneity was evaluated with Cochran’s Q test and horizontal pleiotropy with the MR Egger test.

## Supplemental information titles and legends

**Supplementary Figure 1 – A-B)** Volcano plots showing the protein association profile for incident LOAD, restricted to 10-year follow-up **A)** without *APOE e4* adjustment (model 1) and **B)** with *APOE e4* adjustment (model 2). **C)** Venn diagram for the aptamer overlap between models 1 and 2 for incident AD, restricted to 10-year follow-up.

**Supplementary Figure 2 – A-B)** Volcano plots showing the protein association profile for prevalent LOAD, **A)** without *APOE e4* adjustment (model 1) and **B)** with *APOE e4* adjustment (model 2). **C)** Venn diagram for the aptamer overlap between models 1 and 2 for prevalent LOAD. **D)** Heatmap comparing the effect sizes for models 1 and 2 and both prevalent (odds ratio) and incident LOAD (hazard ratio). Serum protein module membership and *APOE*-dependence (see Methods for definition) are annotated at the top.

**Supplementary Figure 3 –** Spaghetti plots showing the statistical significance of protein associations with **A)** prevalent LOAD and **B)** incident LOAD restricted to 10 year follow-up across the three models, highlighting the 17 *APOE*-dependent proteins (green) defined as those whose association with incident LOAD is attenuated with *APOE* e4 adjustment for incident AD using full follow-up (Fig 2a).

**Supplementary Figure 4 –** Boxplots visualizing protein levels in **A)** AGES serum samples, see Supplementary Table 9 for statistical evaluation and **B)** ACE plasma samples, stratified by APOE-e4 allele carrier and incident LOAD status, for the 17 proteins with APOE-dependent association to incident LOAD in AGES.

**Supplementary Figure 5 – A)** Histogram showing the incident LOAD HR for *APOE-e4* allele count in AGES from Cox proportional hazard regression models adjusted for a single LOAD-associated protein at a time, in addition to age and sex. The dashed line indicates the HR for *APOE-e4* when only adjusting for age and sex. The proteins with the largest effect on the *APOE-e4* HR when included in the model are denoted. **B)** Forest plot showing the incident LOAD HR with 95% confidence intervals for *APOE-e4* when adjusting for each of the 17 *APOE*-dependent model in the Cox proportional hazard regression.

**Supplementary Figure 6 – A-B)** Spaghetti plots showing the statistical significance of protein associations with incident LOAD across three statistical models, adjusting for age and sex, and then additionally either serum **A)** total cholesterol or **B)** LDL cholesterol, and finally APOE e4 carrier status. The LOAD association of the 17 *APOE*-dependent proteins (green) is not attenuated when adjusting for total cholesterol or LDL but only when adjusting for APOE e4 carrier status. **C-H)** Protein associations with serum total cholesterol **(C-E)** and LDL cholesterol **(F-H)**. The density plots show the effect size for the associations between cholesterol levels and all measured proteins stratified by their AD association, adjusting for age and sex (**C** and **F**), and additionally *APOE-e4* carrier status (**D** and **G**). **E)** and **H)** Comparisons of the effect sizes for association with cholesterol levels in the models with (y-axis) and without (x-axis) *APOE-e4* adjustment shows that the effect size is often increased for the *APOE*-dependent proteins (red) after *APOE-e4* adjustment.

**Supplementary Figure 7 –** Reverse MR analysis (excluding the *APOE* locus) for the causal effect of **A)** total cholesterol and **B)** LDL cholesterol on serum levels of the 17 *APOE*-dependent proteins. **C-E)** For each protein, a comparison of the effect sizes for the association with the *APOE-ε4* genotype (x-axis) and **C)** incident LOAD, **D)** total cholesterol and **E)** LDL cholesterol (y-axis). TC, total cholesterol; LDL, low density lipoprotein cholesterol.

**Supplementary Figure 8 –** MR scatterplots for all proteins with more than one genetic instrument and with P<0.05 in the MR analysis for a causal effect of protein levels on AD. Each point represents a genetic instrument (SNP) and shows its effect on serum protein levels (x-axis) and AD (y-axis). The slope of the blue line indicates the inverse variance weighted MR effect.

**Supplementary Figure 9 –** Example MR scatterplots from the reverse MR analysis for a causal effect of AD on serum protein levels, showing discordant direction of effect (inverse variance weighted method indicated by slope of dashed line) when including (left) or excluding (right) APOE genetic variants. Each point represents a genetic instrument (SNP) and shows its effect on AD (x-axis) and serum protein levels (y-axis).

**Supplementary Figure 10 –** A forest plot showing the results for individual SNPs in the reverse MR analysis for a causal effect of AD on serum protein levels, together with the full multi-SNP IVW estimate (red). Plots are shown for the five proteins with FDR<0.1 in the primary reverse MR analysis, excluding variants from the *APOE* locus.

**Supplementary Figure 11 –** Comparison of the effects of *APOE-ε4* (left) versus AD, as evaluated through reverse MR analysis excluding *APOE* variants, (right) on serum protein levels in AGES and two additional cohorts. Results are shown for the four *APOE*-dependent proteins with opposite direction of effect in the two analyses. The AGES results are shown for the full cohort and two age strata.

**Supplementary Table 1 –** Baseline characteristics of the AGES cohort stratified by LOAD status. P values are obtained from t-test for continuous variables, and chi-square test for categorical variables. P and FDR values <0.05 are highlighted in bold.

**Supplementary Table 2 –** Proteins significantly associated with incident LOAD after FDR (Benjamini-Hochberg) correction are displayed. The full Cox proportional hazards regression model (Model 3) was adjusted for baseline age, sex, *APOE-e4* genotype, body mass index, diabetes, education, hypertension, smoking history, and eGFR as denoted below. P and FDR values <0.05 are highlighted in bold. Abbreviations: HR, Hazard ratio; CI, Confidence Interval, Zph: proportional hazard test statistic. Estimates represent difference in LOAD risk per standard deviation increase of aptamer level.

**Supplementary Table 3 –** Proteins significantly associated with incident LOAD with 10-year follow-up after FDR (Benjamini-Hochberg) correction are displayed. The full Cox proportional hazards regression model (Model 3) was adjusted for baseline age, sex, APOE-e4 genotype, body mass index, diabetes, education, hypertension, smoking history, and eGFR. P and FDR values <0.05 are highlighted in bold. Abbreviations: HR, Hazard ratio; CI, Confidence Interval, Zph: proportional hazard test statistic. Estimates represent difference in LOAD risk per standard deviation increase of aptamer level.

**Supplementary Table 4 –** Comparisons of the Cox models with full follow-up vs 10 year follow up using ANOVA. P and FDR values <0.05 are highlighted in bold. Abbreviations: df, degrees of freedom

**Supplementary Table 5 –** Proteins significantly associated with prevalent LOAD after FDR (Benjamini-Hochberg) correction. The full logistic regression model (Model 3) was adjusted for baseline age, sex, APOE-e4, body mass index, diabetes, education, hypertension, smoking history, and eGFR. P and FDR values <0.05 are highlighted in bold. Abbreviations: OR, odds ratio; SE, standard error. Estimates represent difference in LOAD risk per standard deviation increase of aptamer level.

**Supplementary Table 6 –** Replication of the LOAD associated proteins from Supplementary Table 5 in the ACE cohort. Proteins significantly associated with prevalent LOAD after FDR (Benjamini-Hochberg) correction are displayed. Logistic regression models were adjusted for baseline age + sex (Model 1) and additionally for APOE-e4 in Model 2. P and FDR values <0.05 are highlighted in bold. Abbreviations: OR, odds ratio; SE, standard error. Estimates represent difference in LOAD risk per standard deviation increase of aptamer level.

**Supplementary Table 7 –** Results from Gene Set Enrichment Analysis (GSEA) and Overrepresentation Analysis (ORA) of the prevalent LOAD associated proteins against protein-coding gene sets labeled below using the R packages listed below. P values were adjusted for multiple comparisons using FDR (Benjamini Hochberg). Abbreviations: ES, Enrichment score; NES, Enrichment score normalized

**Supplementary Table 8 –** Gene Set Enrichment Analysis and Overrepresentation Analysis of incident LOAD associated proteins in AGES against protein-coding gene sets labeled below using the R packages listed below. P values were adjusted for multiple comparisons using FDR (Benjamini Hochberg). P and FDR values <0.05 are highlighted in bold. Abbreviations: ES, Enrichment score; NES, Enrichment score normalized; size of the pathway after removing missing genes; leadingEdge, vector of genes that drive the enrichment.

**Supplementary Table 9 –** Replication of the incident LOAD associated proteins from Supplementary Table 2 in the ACE cohort. Proteins significantly associated with incident LOAD after FDR (Benjamini-Hochberg) correction are displayed. Linear and Cox proportional regression models were adjusted for baseline age, sex (Model 1) and additionally for APOE-e4 genotype in Model 2. P and FDR values <0.05 are highlighted in bold. Abbreviations: HR, Hazard ratio; CI, Confidence Interval, Zph: proportional hazard test statistic. Estimates represent difference in LOAD risk per standard deviation increase of aptamer level.

**Supplementary Table 10 –** First-degree protein partners of the APOE-dependent proteins from the InWeb database (Li et al. (2016), Nature Methods 2016 14:1 14, 61–64. 10.1038/nmeth.4083). Abbreviations: CS, confidence score.

**Supplementary Table 11 –** Gene Set Enrichment Analysis and Overrepresentation Analysis of the first-degree protein partners of the APOE-dependent LOAD associated proteins against protein-coding gene sets labeled below using the R packages listed below. P values were adjusted for multiple comparisons using FDR (Benjamini Hochberg).

**Supplementary Table 12 –** Forward MR analysis examining the causal effects of the protein partners of the AGES-defined APOE-dependent LOAD-associated proteins on AD. P and FDR values <0.05 are highlighted in bold. Abbreviations: OR, odds ratio; SE, standard error; eggerPass, passes Egger pleiotropy sensitivity analysis; wmPass, passes weighted median sensitivity analysis

**Supplementary Table 13 –** Reverse MR analysis examining the causal effects of blood cholesterol (total and LDL) on the AGES-defined APOE-dependent LOAD-associated proteins. P and FDR values <0.05 are highlighted in bold. Abbreviations: OR, odds ratio; SE, standard error.

**Supplementary Table 14 –** Overview of the genetic instruments and their associations with serum levels in AGES

**Supplementary Table 15 –** Baseline characteristics of the ACE cohort stratified by LOAD status. P values are obtained from t-test for continuous variables, and chi-square test for categorical variables. P and FDR values <0.05 are highlighted in bold.

**Supplementary Table 16 –** Forward MR analysis examining the causal effects of all LOAD-associated proteins in AGES on AD. P and FDR values <0.05 are highlighted in bold. Abbreviations: OR, odds ratio; SE, standard error; eggerPass, passes Egger pleiotropy sensitivity analysis; wmPass, passes weighted median sensitivity analysis

**Supplementary Table 17 –** Reverse MR analysis examining the causal effects of AD or its genetic liability on all LOAD-associated proteins in AGES. The primary analysis was performed without the APOE locus included, but results including the APOE locus are also shown. P and FDR values <0.05 are highlighted in bold. Abbreviations: OR, odds ratio; SE, standard error; Q_pval, Cochran’s Q p-value for instrument heterogeneity

**Supplementary Table 18 –** Replication of reverse MR analysis results in data from Ferkingstad et al. and Sun et al, examining the causal effects of AD on all AGES-defined LOAD-associated proteins. The analysis was performed with and without the APOE locus included for instrument selection. One protein, ARL2, was not available in the Sun et al. data. P and FDR values <0.05 are highlighted in bold. Abbreviations: SE, standard error.

**Supplementary Table 19 –** Proteins associated with prevalent or incident LOAD in AGES that are also associated with AD in either CSF or brain. AGES results are shown for all outcomes and models from which significant proteins were obtained from. P and FDR values <0.05 are highlighted in bold. Abbreviations: HR, Hazard ratio; CI, Confidence Interval; OR, Odds ratio; SE, Standard error.

**Supplementary Table 20 –** Protein association with AD in brain when adjusting for APOE-e4 count (0,1,2) in multinominal logistic regression. Data was obtained from Johnson et al, Nature Neuroscience 2022. Proteins with any association to LOAD in AGES and available data are included in the table, and the HR for incident LOAD in Model 1 and Model 2 are shown for comparison and used to evaluate directional consistency. P and FDR values <0.05 are highlighted in bold. Abbreviations: OR, Odds ratio; SE, Standard error; APOE, APOE-dependent proteins in AGES.

**Supplementary Table 21 –** Serum, CSF and brain protein module membership for all proteins measured in serum in the AGES study. CSF modules were obtained from an extended analysis compared to Dammer et al, Alzheimer’s Research & Therapy 2022 (manuscript in preparation), and brain modules from Johnson et al., Nature Neuroscience 2022. The same protein can occur in more than one module when measured by more than one aptamer (serum and CSF), by different platforms (CSF) or when different peptides are quantified by MS (CSF, brain)

**Supplementary Table 22 –** Overlaps between protein modules (defined from serum, CSF or brain) and proteins associated with AD (in serum, protein or brain) tested by Fisher’s exact test. Serum modules were additionally tested for overlap with proteins detected/measured in CSF and brain, irrespective of their AD association. FDR was calculated using Benjamini-Hochberg. P and FDR values <0.05 are highlighted in bold. Abbreviations: OR, odds ratio; CI, Confidence Interval.

## References

1. Gatz, M. et al. Role of genes and environments for explaining Alzheimer disease. Arch Gen Psychiatry 63, 168–174 (2006).

2. Reitz, C., Rogaeva, E. & Beecham, G. W. Late-onset vs nonmendelian early-onset Alzheimer disease: A distinction without a difference? Neurol Genet 6, (2020).

3. Rajan, K. B. et al. Population Estimate of People with Clinical AD and Mild Cognitive Impairment in the United States (2020–2060). Alzheimers Dement 17, 1966 (2021).

4. van Dyck, C. H. et al. Lecanemab in Early Alzheimer’s Disease. New England Journal of Medicine 388, 9–21 (2023).

5. Mintun, M. A. et al. Donanemab in Early Alzheimer’s Disease. New England Journal of Medicine 384, 1691–1704 (2021).

6. Sattlecker, M. et al. Longitudinal Protein Changes in Blood Plasma Associated with the Rate of Cognitive Decline in Alzheimer’s Disease. Journal of Alzheimer’s Disease 49, 1105–1114 (2016).

7. Kiddle, S. J. et al. Candidate Blood Proteome Markers of Alzheimer’s Disease Onset and Progression: A Systematic Review and Replication Study. Journal of Alzheimer’s Disease 38, 515–531 (2014).

8. Sattlecker, M. et al. Alzheimer’s disease biomarker discovery using SOMAscan multiplexed protein technology. Alzheimer’s & Dementia 10, 724–734 (2014).

9. O’Bryant, S. E. et al. A Serum Protein-Based Algorithm for the Detection of Alzheimer’s Disease. Arch Neurol 67, 1077 (2010).

10. Ijsselstijn, L. et al. Serum levels of pregnancy zone protein are elevated in presymptomatic alzheimer’s disease. J Proteome Res 10, 4902–4910 (2011).

11. Ray, S. et al. Classification and prediction of clinical Alzheimer’s diagnosis based on plasma signaling proteins. Nature Medicine 2007 13:11 13, 1359–1362 (2007).

12. Henkel, A. W. et al. Multidimensional plasma protein separation technique for identification of potential Alzheimer’s disease plasma biomarkers: A pilot study. J Neural Transm 119, 779–788 (2012).

13. Choi, J., Malakowsky, C. A., Talent, J. M., Conrad, C. C. & Gracy, R. W. Identification of oxidized plasma proteins in Alzheimer’s disease. Biochem Biophys Res Commun 293, 1566–1570 (2002).

14. Cutler, P. et al. Proteomic identification and early validation of complement 1 inhibitor and pigment epithelium-derived factor: Two novel biomarkers of Alzheimer’s disease in human plasma. Proteomics Clin Appl 2, 467–477 (2008).

15. Hye, A. et al. Proteome-based plasma biomarkers for Alzheimer’s disease. Brain 129, 3042–3050 (2006).

16. Doecke, J. D. et al. Blood-Based Protein Biomarkers for Diagnosis of Alzheimer Disease. Arch Neurol 69, 1318 (2012).

17. Kiddle, S. J. et al. Candidate Blood Proteome Markers of Alzheimer’s Disease Onset and Progression: A Systematic Review and Replication Study. Journal of Alzheimer’s Disease 38, 515–531 (2014).

18. Walker, K. A. et al. Large-scale plasma proteomic analysis identifies proteins and pathways associated with dementia risk. Nature Aging 2021 1:5 1, 473–489 (2021).

19. Bellenguez, C. et al. New insights into the genetic etiology of Alzheimer’s disease and related dementias. Nature Genetics 2022 1–25 (2022) doi:10.1038/s41588-022-01024-z.

20. Khani, M., Gibbons, E., Bras, J. & Guerreiro, R. Challenge accepted: uncovering the role of rare genetic variants in Alzheimer’s disease. Mol Neurodegener 17, 3 (2022).

21. Frisoni, G. B. et al. The prevalence of apoE-ε4 in Alzheimer’s disease is age dependent. J Neurol Neurosurg Psychiatry 65, 103 (1998).

22. Gharbi-Meliani, A. et al. The association of APOE ε4 with cognitive function over the adult life course and incidence of dementia: 201years follow-up of the Whitehall II study. Alzheimers Res Ther 13, (2021).

23. Corder, E. H. et al. Gene Dose of Apolipoprotein E Type 4 Allele and the Risk of Alzheimer’s Disease in Late Onset Families. Science (1979) 261, 921–923 (1993).

24. Gudjonsson, A. et al. A genome-wide association study of serum proteins reveals shared loci with common diseases. Nature Communications 2022 13:1 13, 1–13 (2022).

25. Sun, B. B. et al. Genomic atlas of the human plasma proteome. Nature 2018 558:7708 558, 73–79 (2018).

26. Emilsson, V. et al. Co-regulatory networks of human serum proteins link genetics to disease. Science 361, (2018).

27. Emilsson, V. et al. Coding and regulatory variants are associated with serum protein levels and disease. Nature Communications 2022 13:1 13, 1–11 (2022).

28. Cruchaga, C. et al. Proteogenomic analysis of human cerebrospinal fluid identifies neurologically relevant regulation and informs causal proteins for Alzheimer’s disease. Res Sq (2023) doi:10.21203/rs.3.rs-2814616/v1.

29. Yang, C. et al. Genomic atlas of the proteome from brain, CSF and plasma prioritizes proteins implicated in neurological disorders. Nat Neurosci 24, 1302–1312 (2021).

30. Harris, T. B. et al. Age, Gene/Environment Susceptibility – Reykjavik Study: Multidisciplinary Applied Phenomics. Am J Epidemiol 165, 1076 (2007).

31. Uhlén, M. et al. Proteomics. Tissue-based map of the human proteome. Science 347, (2015).

32. Li, T. et al. A scored human protein–protein interaction network to catalyze genomic interpretation. Nature Methods 2016 14:1 14, 61–64 (2016).

33. Postmus, I. et al. Pharmacogenetic meta-analysis of genome-wide association studies of LDL cholesterol response to statins. Nat Commun 5, (2014).

34. Marucci, G. et al. Efficacy of acetylcholinesterase inhibitors in Alzheimer’s disease. Neuropharmacology 190, (2021).

35. Ferkingstad, E. et al. Large-scale integration of the plasma proteome with genetics and disease. Nat Genet 53, 1712–1721 (2021).

36. Dammer, E. B. et al. Proteomic Network Analysis of Alzheimer’s Disease Cerebrospinal Fluid Reveals Alterations Associated with APOE 14 Genotype and Atomoxetine Treatment. medRxiv 2023.10.29.23297651 (2023) doi:10.1101/2023.10.29.23297651.

37. Johnson, E. C. B. et al. Large-scale deep multi-layer analysis of Alzheimer’s disease brain reveals strong proteomic disease-related changes not observed at the RNA level. Nature Neuroscience 2022 25:2 25, 213–225 (2022).

38. Dammer, E. B. et al. Multi-platform proteomic analysis of Alzheimer’s disease cerebrospinal fluid and plasma reveals network biomarkers associated with proteostasis and the matrisome. Alzheimers Res Ther 14, (2022).

39. Porter, T. et al. Cognitive gene risk profile for the prediction of cognitive decline in presymptomatic Alzheimer’s disease. Pers Med Psychiatry 7–8, 14–20 (2018).

40. Zhou, M. et al. Targeted mass spectrometry to quantify brain-derived cerebrospinal fluid biomarkers in Alzheimer’s disease. Clin Proteomics 17, 1–14 (2020).

41. Lourenço, F. C. et al. Netrin-1 interacts with amyloid precursor protein and regulates amyloid-beta production. Cell Death Differ 16, 655–663 (2009).

42. Zetterberg, H. Neurofilament Light: A Dynamic Cross-Disease Fluid Biomarker for Neurodegeneration. Neuron 91, 1–3 (2016).

43. Graham, N. S. N. et al. Axonal marker neurofilament light predicts long-term outcomes and progressive neurodegeneration after traumatic brain injury. Sci Transl Med 13, (2021).

44. Wolters, F. et al. Von Willebrand Factor and the Risk of Dementia: A Population-Based Study (P1.092). Neurology 86, (2016).

45. Johnson, E. C. B. et al. Cerebrospinal fluid proteomics define the natural history of autosomal dominant Alzheimer’s disease. Nat Med (2023) doi:10.1038/s41591-023-02476-4.

46. Sattlecker, M. et al. Alzheimer’s disease biomarker discovery using SOMAscan multiplexed protein technology. Alzheimer’s & Dementia 10, 724–734 (2014).

47. Lindbohm, J. V. et al. Plasma proteins, cognitive decline, and 20-year risk of dementia in the Whitehall II and Atherosclerosis Risk in Communities studies. Alzheimer’s & Dementia 18, 612 (2022).

48. Eldjarn, G. H. et al. Large-scale plasma proteomics comparisons through genetics and disease associations. Nature 622, 348–358 (2023).

49. Sebastiani, P. et al. A serum protein signature of APOE genotypes in centenarians. Aging Cell 18, e13023 (2019).

50. Uhlén, M. et al. Proteomics. Tissue-based map of the human proteome. Science 347, (2015).

51. Buniello, A. et al. The NHGRI-EBI GWAS Catalog of published genome-wide association studies, targeted arrays and summary statistics 2019. Nucleic Acids Res 47, D1005–D1012 (2019).

52. Jun, G. et al. A NOVEL ALZHEIMER DISEASE LOCUS LOCATED NEAR THE GENE ENCODING TAU PROTEIN. Mol Psychiatry 21, 108 (2016).

53. Okbay, A. et al. Polygenic prediction of educational attainment within and between families from genome-wide association analyses in 3 million individuals. Nat Genet 54, 437–449 (2022).

54. Brouwer, R. M. et al. Genetic variants associated with longitudinal changes in brain structure across the lifespan. Nature Neuroscience 2022 25:4 25, 421–432 (2022).

55. Davies, G. et al. Study of 300,486 individuals identifies 148 independent genetic loci influencing general cognitive function. Nat Commun 9, (2018).

56. Wang, H. et al. Genome-wide interaction analysis of pathological hallmarks in Alzheimer’s disease. Neurobiol Aging 93, 61–68 (2020).

57. Tin, A. et al. Proteomic Analysis Identifies Circulating Proteins Associated With Plasma Amyloid β and Incident Dementia. Biological Psychiatry Global Open Science (2022) doi:10.1016/J.BPSGOS.2022.04.005.

58. Tanzi, R. E. et al. Amyloid beta protein gene: cDNA, mRNA distribution, and genetic linkage near the Alzheimer locus. Science 235, 880–884 (1987).

59. Bai, Z. et al. Distinctive RNA Expression Profiles in Blood Associated with Alzheimer’s Disease after Accounting for White Matter Hyperintensities. Alzheimer Dis Assoc Disord 28, 226–233 (2014).

60. Wang, D. et al. Cardiotrophin-1 (CTF1) ameliorates glucose-uptake defects and improves memory and learning deficits in a transgenic mouse model of Alzheimer’s disease. Pharmacol Biochem Behav 107, 48–57 (2013).

61. Rayaprolu, S. et al. Flow-cytometric microglial sorting coupled with quantitative proteomics identifies moesin as a highly-abundant microglial protein with relevance to Alzheimer’s disease. Mol Neurodegener 15, 28 (2020).

62. Chen, H.-H. et al. Genetically regulated expression in late-onset Alzheimer’s disease implicates risk genes within known and novel loci. Transl Psychiatry 11, 618 (2021).

63. Sügis, E. et al. HENA, heterogeneous network-based data set for Alzheimer’s disease. Sci Data 6, 151 (2019).

64. Zetterberg, H. et al. Association of Cerebrospinal Fluid Neurofilament Light Concentration With Alzheimer Disease Progression. JAMA Neurol 73, 60 (2016).

65. Bernardini, S. et al. Glutathione S-Transferase P1 *C Allelic Variant Increases Susceptibility for Late-Onset Alzheimer Disease: Association Study and Relationship with Apolipoprotein E ε4 Allele. Clin Chem 51, 944–951 (2005).

66. Pinhel, M. A. S. et al. Glutathione S-transferase variants increase susceptibility for late-onset Alzheimer’s disease: association study and relationship with apolipoprotein E 14 allele. Clin Chem Lab Med 46, (2008).

67. Sigurdsson, S. et al. Incidence of Brain Infarcts, Cognitive Change, and Risk of Dementia in the General Population: The AGES-Reykjavik Study (Age Gene/Environment Susceptibility-Reykjavik Study). Stroke 48, 2353–2360 (2017).

68. Jørgensen, L. M., El Kholy, K., Damkjær, K., Deis, A. & Schroll, M. »RAI«1-Et internationalt system til vurdering af beboere på plejehjem. Ugeskr Laeger 159, 6371–6376 (1997).

69. Gudnason V, S. J. S. L. H. S. S. G. Association of apolipoprotein E polymorphism with plasma levels of high density lipoprotein and lipoprotein(a), and effect of diet in healthy men and women. NUTRITION METABOLISM AND CARDIOVASCULAR DISEASES 3, 136–141 (1993).

70. Levey, A., Greene, T., Kusek, J. & Beck, G. A simplified equation to predict glomerular filtration rate from serum creatinine. Journal of the American Society of Nephrology 11, 155A (2000).

71. Gudmundsdottir, V. et al. Circulating Protein Signatures and Causal Candidates for Type 2 Diabetes. Diabetes 69, 1843 (2020).

72. Lamb, J. R., Jennings, L. L., Gudmundsdottir, V., Gudnason, V. & Emilsson, V. It’s in Our Blood: A Glimpse of Personalized Medicine. Trends Mol Med 27, 20–30 (2021).

73. Gold, L. et al. Aptamer-Based Multiplexed Proteomic Technology for Biomarker Discovery. doi:10.1371/journal.pone.0015004.

74. Tuck, M. K. et al. Standard operating procedures for serum and plasma collection: Early detection research network consensus statement standard operating procedure integration working group. J Proteome Res 8, 113–117 (2009).

75. Jessen, F. et al. A conceptual framework for research on subjective cognitive decline in preclinical Alzheimer’s disease. Alzheimer’s & Dementia 10, 844–852 (2014).

76. Lopez, O. L. et al. Risk Factors for Mild Cognitive Impairment in the Cardiovascular Health Study Cognition Study. Arch Neurol 60, 1394 (2003).

77. Petersen, R. C. et al. Mild cognitive impairment: a concept in evolution. J Intern Med 275, 214–228 (2014).

78. Petersen, R. C. et al. Mild Cognitive Impairment. Arch Neurol 56, 303 (1999).

79. Jack, C. R. et al. NIA-AA Research Framework: Toward a biological definition of Alzheimer’s disease. Alzheimer’s & Dementia 14, 535–562 (2018).

80. Orellana, A. et al. Establishing In-House Cutoffs of CSF Alzheimer’s Disease Biomarkers for the AT(N) Stratification of the Alzheimer Center Barcelona Cohort. Int J Mol Sci 23, 6891 (2022).

81. Rodriguez-Gomez, O. et al. FACEHBI: A PROSPECTIVE STUDY OF RISK FACTORS, BIOMARKERS AND COGNITION IN A COHORT OF INDIVIDUALS WITH SUBJECTIVE COGNITIVE DECLINE. STUDY RATIONALE AND RESEARCH PROTOCOLS. J Prev Alzheimers Dis 1–9 (2016) doi:10.14283/jpad.2016.122.

82. Moreno-Grau, S. et al. Genome-wide association analysis of dementia and its clinical endophenotypes reveal novel loci associated with Alzheimer’s disease and three causality networks: The GR@ACE project. Alzheimer’s & Dementia 15, 1333–1347 (2019).

83. Vanderstichele, H. et al. Standardization of preanalytical aspects of cerebrospinal fluid biomarker testing for Alzheimer’s disease diagnosis: A consensus paper from the Alzheimer’s Biomarkers Standardization Initiative. Alzheimer’s & Dementia 8, 65–73 (2012).

84. Candia, J., Daya, G. N., Tanaka, T., Ferrucci, L. & Walker, K. A. Assessment of variability in the plasma 7k SomaScan proteomics assay. Sci Rep 12, 17147 (2022).

85. Therneau, T., Crowson, C. & Clinic, M. Using Time Dependent Covariates and Time Dependent Coefficients in the Cox Model. (2014).

86. Gottesman, R. F. et al. Associations Between Midlife Vascular Risk Factors and 25-Year Incident Dementia in the Atherosclerosis Risk in Communities (ARIC) Cohort. JAMA Neurol 74, 1246 (2017).

87. Yu, G., Wang, L. G., Han, Y. & He, Q. Y. clusterProfiler: an R Package for Comparing Biological Themes Among Gene Clusters. OMICS 16, 284 (2012).

88. Korotkevich, G. et al. Fast gene set enrichment analysis. bioRxiv 060012 (2021) doi:10.1101/060012.

89. Subramanian, A. et al. Gene set enrichment analysis: A knowledge-based approach for interpreting genome-wide expression profiles. Proc Natl Acad Sci U S A 102, 15545–15550 (2005).

90. Aguet, F. et al. The GTEx Consortium atlas of genetic regulatory effects across human tissues. Science 369, 1318 (2020).

91. Beach, T. G. et al. Arizona Study of Aging and Neurodegenerative Disorders and Brain and Body Donation Program. Neuropathology 35, 354–389 (2015).

92. Bennett, D. A. et al. Religious Orders Study and Rush Memory and Aging Project. Journal of Alzheimer’s Disease 64, S161–S189 (2018).

93. Jack, C. R. et al. NIA-AA Research Framework: Toward a biological definition of Alzheimer’s disease. Alzheimer’s & Dementia 14, 535–562 (2018).

94. Burgess, S., Dudbridge, F. & Thompson, S. G. Combining information on multiple instrumental variables in Mendelian randomization: comparison of allele score and summarized data methods. Stat Med 35, 1880 (2016).

