## Supplementary meterial for "Serum proteomics reveals *APOE* dependent and independent protein signatures in Alzheimer’s disease"

1	Icelandic Heart Association, Kopavogur, 200, Iceland
2	Faculty of Medicine, University of Iceland, Reykjavik, 101, Iceland
3	Goizueta Alzheimer’s Disease Research Center, Emory University School of Medicine, Atlanta, 30329,GA, USA
4	Department of Neurology, Emory University School of Medicine, Atlanta, 30329, GA, USA
5	Department of Biochemistry, Emory University School of Medicine, Atlanta, 30329, GA, USA
6	Research Center and Memory Clinic. Ace Alzheimer Center Barcelona – Universitat Internacional de Catalunya, Barcelona,
 08028, Spain, Barcelona.
7	CIBERNED, Network Center for Biomedical Research in Neurodegenerative Diseases, National Institute of Health Carlos III,
 Madrid, 28029, Spain.
8	Novartis Institutes for Biomedical Research, Cambridge, 02139, MA, USA
9	Novartis Institutes for Biomedical Research, San Diego, 10675, CA, USA
10 Laboratory of Epidemiology and Population Sciences, Intramural Research Program, National Institute
 on Aging, Bethesda, 20892, MD, USA

Supplementary Figure 1	7

Supplementary Figure 2	8

Supplementary Figure 3	9

Supplementary Figure 4	11

Supplementary Figure 6	13

Supplementary Figure 7	14

Supplementary Figure 8	15

Supplementary Figure 9	16

Supplementary Figure 10	17

Supplementary Figure 11	18

### Supplementary Notes

#### Supplementary Note 1 – Protein associations with incident LOAD limited to 10-year follow up time

To account for hazard ratio variability that can arise with lengthy follow-up time, secondary analyses were implemented with a 10-year follow-up cut-off, during which 432 individuals developed LOAD. Here, a total of 106 proteins were associated (FDR < 0.05) with incident LOAD in *model 1* and 21 of them were specific to the shorter follow-up analysis compared to the primary analysis using full follow-up. 34 proteins were significant in *model 2* when adjusting for *APOE*-ε4, of which 10 were specific to this analysis. No proteins were significant in *model 3* (Supplementary Table 2). For the proteins specific to the shorter follow-up analysis, we tested whether their associations with LOAD were time-dependent. Here we found that the effect was limited to the first 10 years (ANOVA P<0.05) (Supplementary Table 4). Thus, these proteins yield a separation of cases and non-LOAD individuals during the first 10 years of follow-up which then diminishes over time, suggesting they may reflect processes that take place closer to the LOAD diagnosis. These included proteins with established roles in the CNS (MAPT, CARTPT, OSTN, PCDHGC5, LRRTM4) and the immune system (DEFB103B, CD70, TNFSF10 and TNFRSF14). Intriguingly, the tau protein, as measured by the MAPT (Microtubule-associated protein tau) aptamer, showed a protective association with incident LOAD (HR = 0.83, 95% CI: 0.45-0.92, P = 2.2e-4), although it is unclear which tau isoform is targeted by the aptamer.

#### Supplementary Note 2 – Protein associations with prevalent LOAD.

By virtue of the AGES cohort design, we were able to study LOAD on a longitudinal level as well as on a cross-sectional level when considering prevalent LOAD cases (n = 167, Figure 1). A cross-sectional comparison of the serum protein profile of individuals with prevalent LOAD to controls using a logistic regression analysis resulted in the detection of eight unique statistically significant (FDR < 0.05) proteins for model 1 (S100A13, TBCA, ARL2, IRF6, EHMT2, CKAP4, LRRN1 and CLEC3B) (Supplementary Table 5). The abundances of all these proteins were negatively associated with LOAD status except for LRRN1, with odds ratios (ORs) ranging from 0.65 per standard deviation elevation for TBCA to 1.36 for LRRN1 (Figure 1A). In model 2, four significant (FDR < 0.05) proteins were identified, including ERAP1 which was new compared to model 1 (Figure 1B-C). ORs ranged from 0.71 for ERAP1 to 0.66 for EHMT2 (Figure 1B, Supplementary Table 5). Model 3 attenuated some of the observed associations and resulted in one additional significant (FDR < 0.05) association for SPINK9 (OR = 0.68) (Supplementary Table 5). Of the ten proteins, three (CKAP4, S100A13 and SPINK9) were also associated (FDR < 0.05) with prevalent LOAD compared to individuals with MCI in the ACE cohort (Supplementary Table 6 and 14). Intriguingly only SPINK9 was directionally consistent between the two cohorts.

The low number of associations detected in the cross-sectional analysis may reflect the smaller number of prevalent compared to incident cases, differences between the prevalent and incident LOAD populations, including a potential mortality bias among the prevalent LOAD cases, or dynamic protein fluctuations along the disease continuum.

To investigate the potential processes reflected by the protein profile associated with prevalent LOAD in AGES, we performed a gene set enrichment analysis (GSEA), which showed an enrichment of transmembrane receptor protein tyrosine kinase activity, actin folding and formation of tubulin folding intermediates (Supplementary Table 7). Over-Representation Analysis (ORA) looking exclusively at the proteins that were significantly associated with prevalent LOAD (model 1) furthermore revealed ties to lysine degradation, histone modifications, chaperonin tubulin folding and subcutaneous adipose tissue (Supplementary Table 7).

#### Supplementary Note 3 - Network analysis of APOE-dependent proteins

Given that the 17 *APOE*-dependent proteins may be a readout of mechanisms that mediate *APOE-*ε*4* risk on LOAD while also being affected in the opposite direction by general LOAD liability, understanding which biological processes and systems these proteins reflect is critical. To further understand the biological context and function of the 17 *APOE*-dependent proteins defined in our study, we investigated their protein-protein interaction (PPI) partners, as individual proteins are rarely uniquely responsible for phenotypic traits or biological functions. Furthermore, they could shed light on how the LOAD associated serum proteins are related to the disease. We detected 365 first-degree protein partners via the InWeb^1^ database of measured and inferred physical PPIs (Supplementary Table 10). Over-representation analysis of the protein partners resulted in a broad enrichment profile of AD-relevant terms over various enrichment gene sets. Terms related to proteasomal regulation, ubiquitination, neurotrophin signaling pathways, and neuronal response- and development were dominant in more than one gene set. Alzheimer-specific terms were also significant such as “Amyloid fibril formation”, “Cellular response to amyloid-beta” and, “Alzheimer’s disease” (Figure 3, Supplementary Table 11). Thus, we demonstrate that the serum proteins with an *APOE*-dependent association to LOAD in AGES are known to physically interact with proteins directly involved in processes reflecting LOAD pathogenesis in the brain.

As only two (C1orf56 and GSTM1) of the 17 APOE-dependent proteins could be tested for a causal role in LOAD in the forward MR analysis due to lack of available cis-instruments, we subjected their PPI partners to a MR analysis. Of the PPI partners, 17 had a cis-pQTL in AGES and could be tested for causality. We detected two proteins with support for causality, APP which alongside our previously described observational and MR associations, emerged as a protein partner for NEFL (MR OR = 0.76, FDR = 0.005) and Mitogen-Activated Protein Kinase 3 (MAPK3, MR OR = 1.43, FDR = 0.009), which is a protein partner for MSN (Supplementary Table 12).

#### Supplementary Note 4 – The effect of cholesterol levels on the association between *APOE*-dependent proteins and LOAD

Due to the well-established relationship between APOE and cholesterol^2^ we investigated the potential effect that serum lipid levels might have on the association between LOAD and the 17 *APOE-*dependent proteins. After adjusting for total and LDL cholesterol on the Cox proportional hazard models, we found that the protein associations with LOAD were mostly preserved (Supplementary Figure 5A-B). For some proteins, their association to LOAD was even stronger after the adjustment (Supplementary Figure 5A-B). However, we additionally found that the *APOE-*dependent proteins were more strongly associated with total serum cholesterol and LDL cholesterol levels compared to other measured proteins (Supplementary Figure 5C-H). Interestingly, these associations between proteins and cholesterol levels were independent of the *APOE-*ε*4* genotype and similarly as for LOAD, the association was in some cases found to be stronger upon *APOE-*ε*4* adjustment (Supplementary Figure 5C-H). These findings suggest that, while the *APOE*-dependent proteins are in fact linked to cholesterol, this relationship is not the driver of their link to LOAD. At the same time, the *APOE-*ε*4* allele is not the key driver of the association between these proteins and cholesterol.

To further explore the observed relationship between 17 *APOE*-dependent proteins and cholesterol levels, we performed a reverse MR analysis (excluding the *APOE* locus) to investigate the causal effect of total cholesterol and LDL cholesterol on the serum levels of these protein (see Methods). We found significant (FDR < 0.05) support for causal effects of total cholesterol and/or LDL cholesterol on increased levels of 13 of the 17 proteins (Supplementary Figure 6, Supplementary Table 13), including all the 12 downregulated by the *APOE-*ε*4* allele (Figure 2C), even though the allele itself is known to increase cholesterol levels^3,4^. These findings further underscore that the association between the *APOE*-dependent proteins and cholesterol is not explained by *APOE-*ε*4.* Furthermore, these findings suggest that the levels of these proteins are affected by LDL cholesterol levels, raising the question if their LOAD associations may be mediated by cholesterol metabolism in the brain.

#### Supplementary Note 5 – Secondary reverse MR analysis including the *APOE* locus, evaluating the effect of LOAD on serum protein levels

The primary reverse MR analysis investigating the causal effect of LOAD, or its genetic liability, on serum protein levels was performed using AD-associated genetic variants as instruments but excluding variants in the *APOE* locus (see Methods and Results), due to its established pleiotropic effects both at the molecular and phenotypic level. However, we performed a secondary reverse MR analysis including variants in the *APOE* locus as instruments. Here we observed support for reverse causality of LOAD on serum levels of 23 proteins (Supplementary Table 17). However, for almost half (10 of 23) of these proteins, we observed significant heterogeneity between instruments (Cochran’s Q P < 0.05), indicating a major influence of *APOE* variants on the results and a violation of the MR assumptions. An example of this is demonstrated in Supplementary Figure 9, showing scatterplots for the instruments used in the reverse MR analysis with and without variants in the *APOE* locus for two proteins (ARL2 and S100A13), where *APOE* variants dominate the effect in the former. Thus, the reverse MR results that include *APOE* variants seem to mainly reflect the effects of the *APOE* locus itself on serum proteins and not the general genetic liability for LOAD. It is worth mentioning that we did not detect any change in effect for *APOE*-independent proteins in the reverse MR analyses when including or excluding the *APOE* locus (Supplementary Table 17), which was not surprising as per definition these proteins were not affected by *APOE-ε4* adjustment in the observational analysis.

### Supplementary Figures

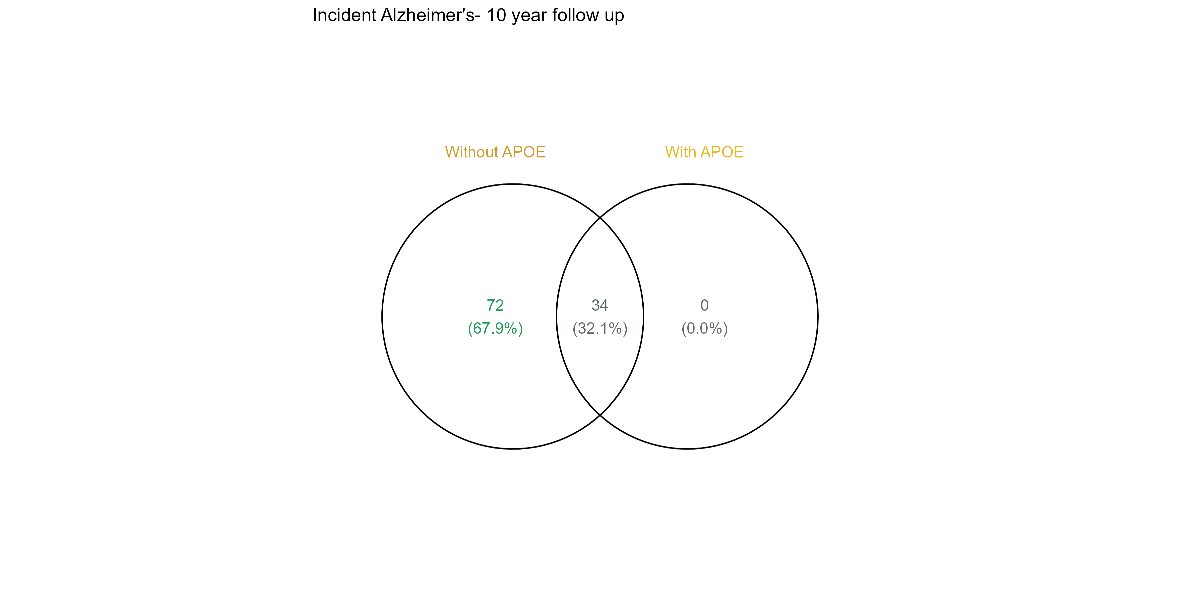

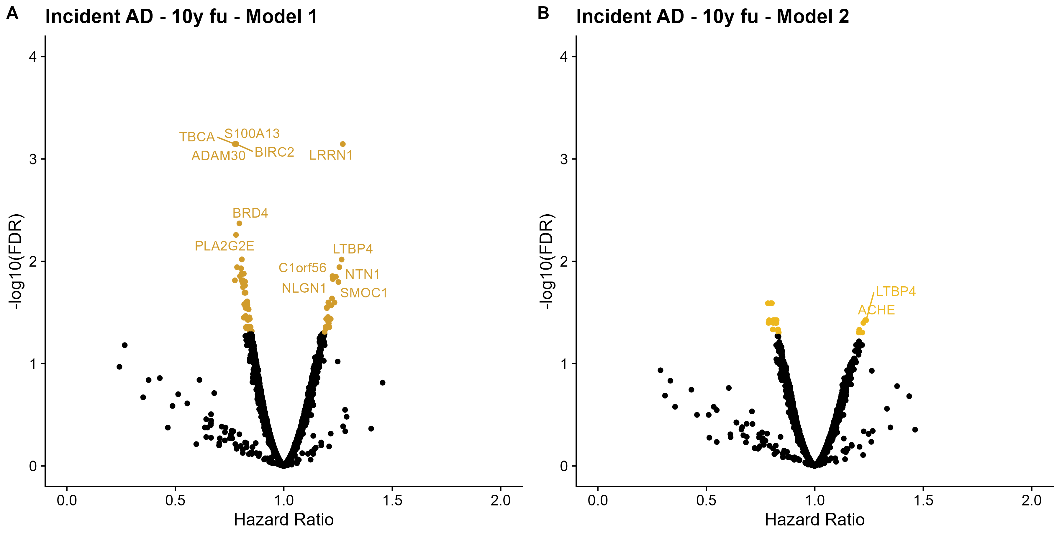

**A	B	C**

Supplementary Figure 1 – **A-B)** Volcano plots showing the protein association profile for incident LOAD, restricted to 10-year follow-up **A)** without *APOE e4* adjustment (model 1) and **B)** with *APOE e4* adjustment (model 2). **C)** Venn diagram for the SOMAmer overlap between models 1 and 2 for incident AD, restricted to 10-year follow-up.

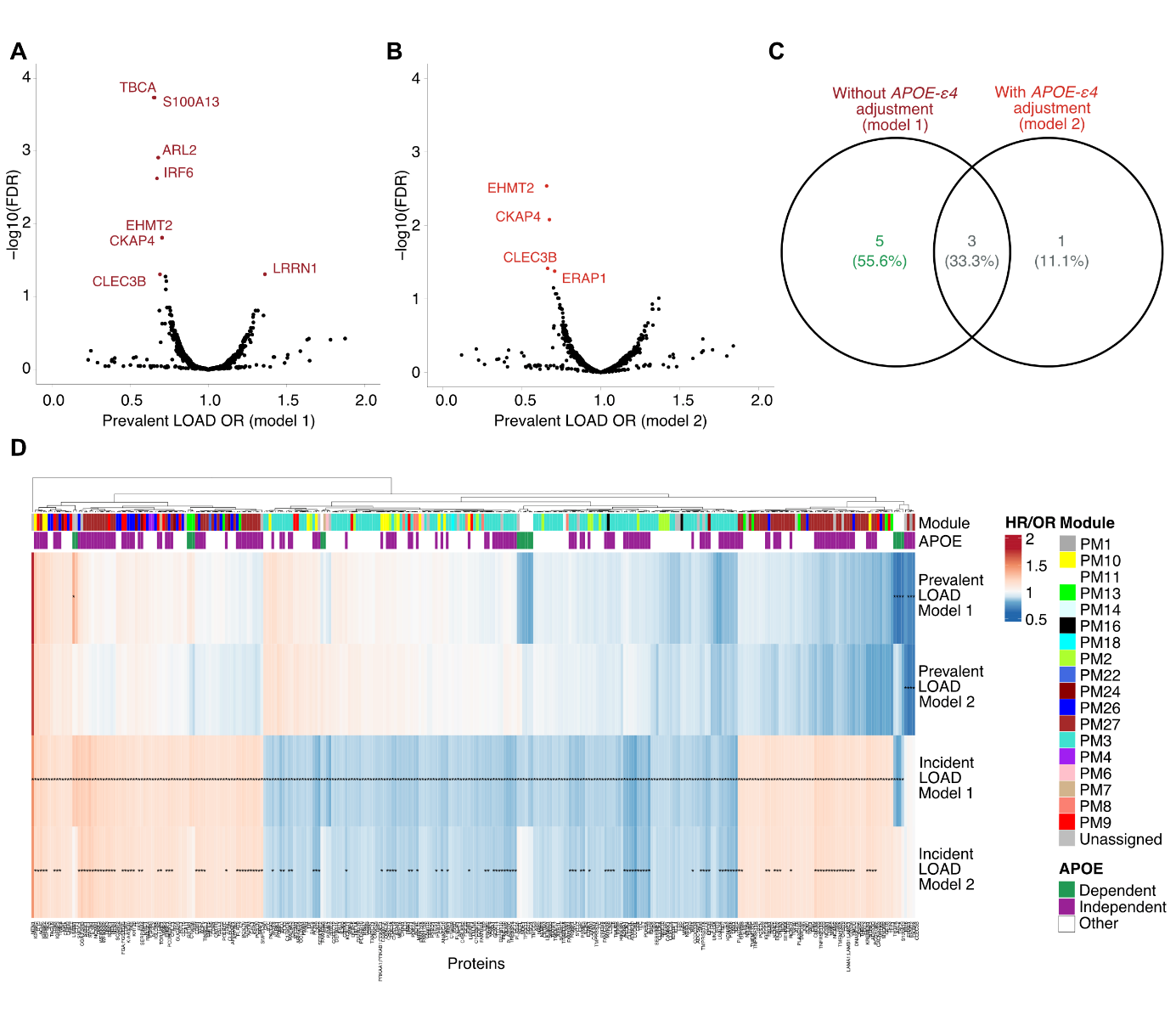

Supplementary Figure 2 – **A-B)** Volcano plots showing the protein association profile for prevalent LOAD, **A)** without *APOE e4* adjustment (model 1) and **B)** with *APOE e4* adjustment (model 2). **C)** Venn diagram for the SOMAmer overlap between models 1 and 2 for prevalent LOAD. **D)** Heatmap comparing the effect sizes for models 1 and 2 and both prevalent (odds ratio) and incident LOAD (hazard ratio). Serum protein module membership and *APOE*-dependence (see Methods for definition) are annotated at the top.

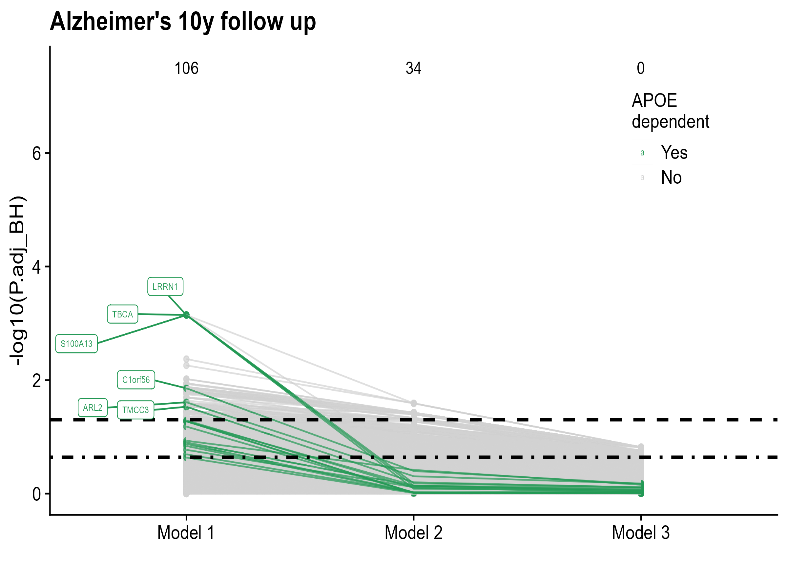

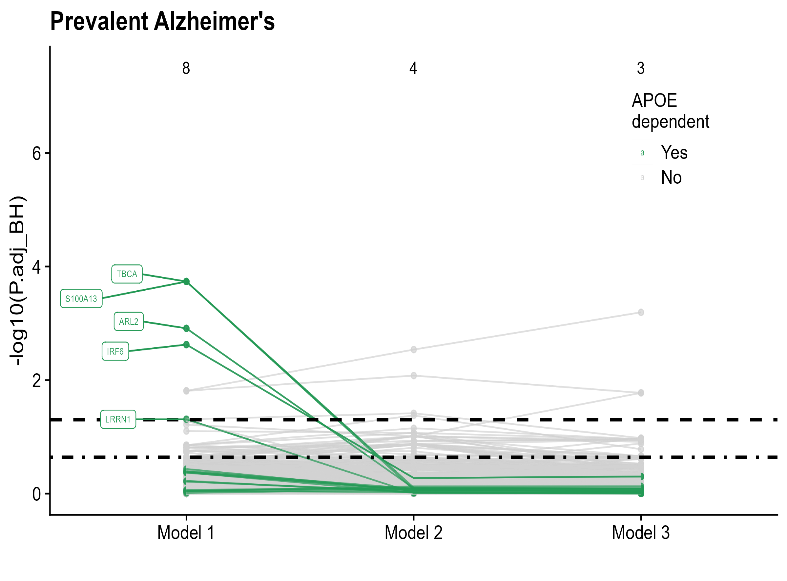

Supplementary Figure 3 – Spaghetti plots showing the statistical significance of protein associations with **A)** prevalent LOAD and **B)** incident LOAD restricted to 10 year follow-up across the three models, highlighting the 17 *APOE*-dependent proteins (green) defined as those whose association with incident LOAD is attenuated with *APOE* e4 adjustment for incident AD using full follow-up (**Fig 2a**).

**A	B**

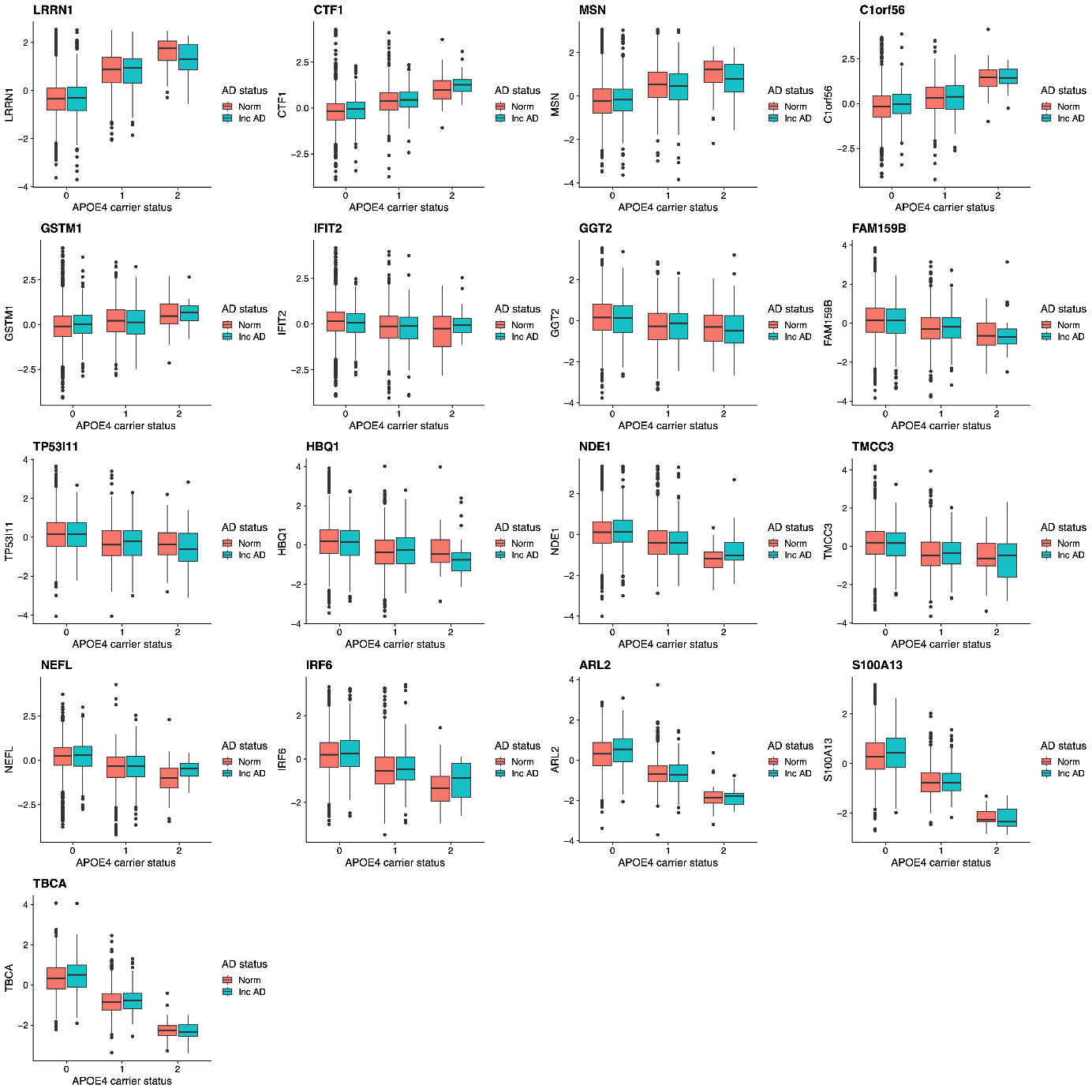

A

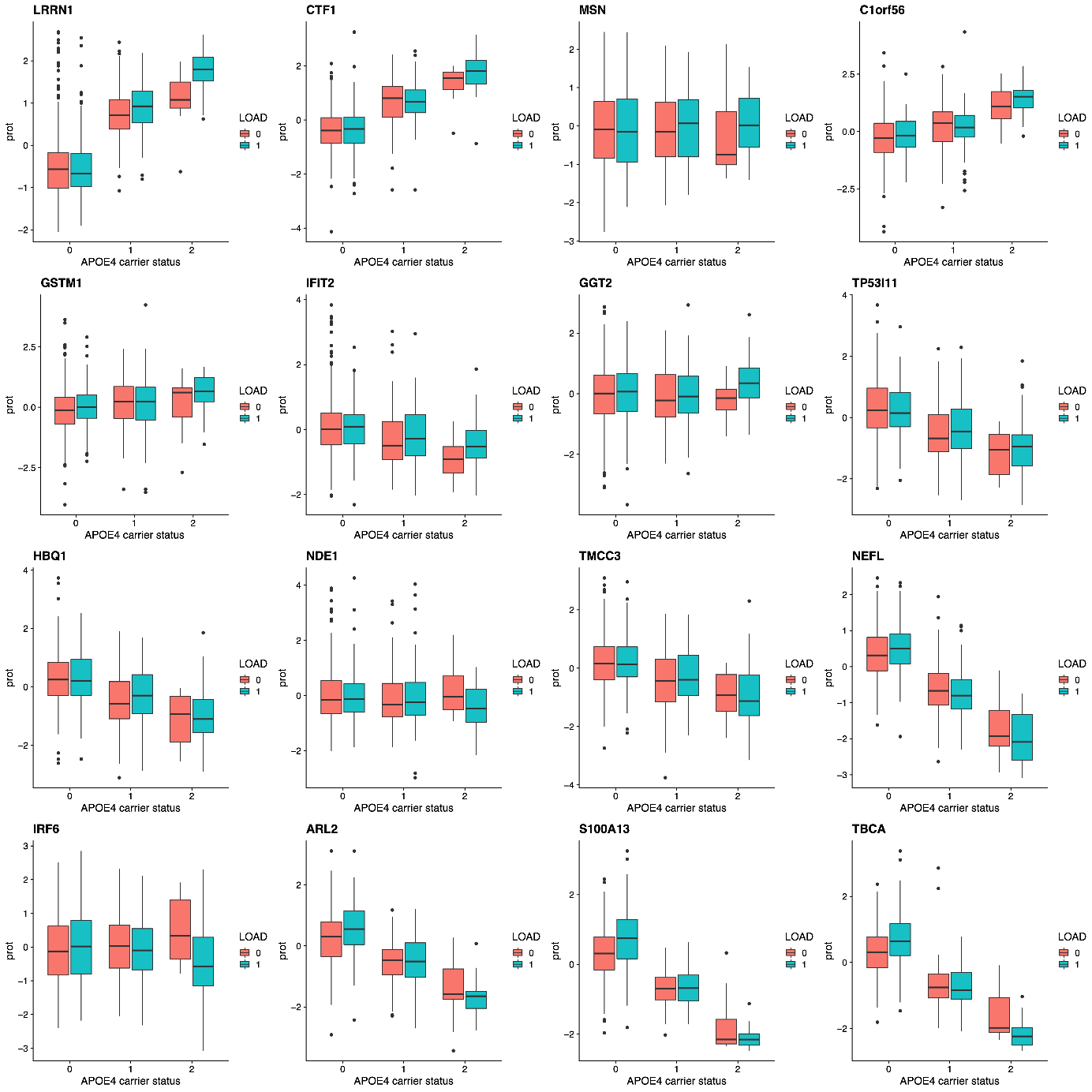

B

Supplementary Figure 4 – Boxplots visualizing protein levels in **A)** AGES serum samples, see Supplementary Table 9 for statistical evaluation and **B)** ACE plasma samples, stratified by APOE-e4 allele carrier and incident LOAD status, for the 17 proteins with APOE-dependent association to incident LOAD in AGES.

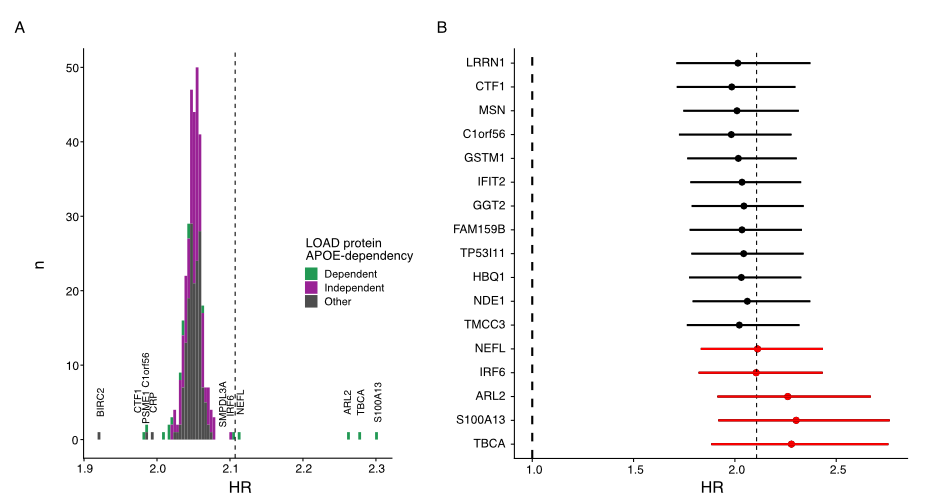

Supplementary Figure 5 – **A)** Histogram showing the incident LOAD HR for *APOE-e4* allele count in AGES from Cox proportional hazard regression models adjusted for a single LOAD-associated protein at a time, in addition to age and sex. The dashed line indicates the HR for *APOE-e4* when only adjusting for age and sex. The proteins with the largest effect on the *APOE-e4* HR when included in the model are denoted. **B)** Forest plot showing the incident LOAD HR with 95% confidence intervals for *APOE-e4* when adjusting for each of the 17 *APOE*-dependent model in the Cox proportional hazard regression.

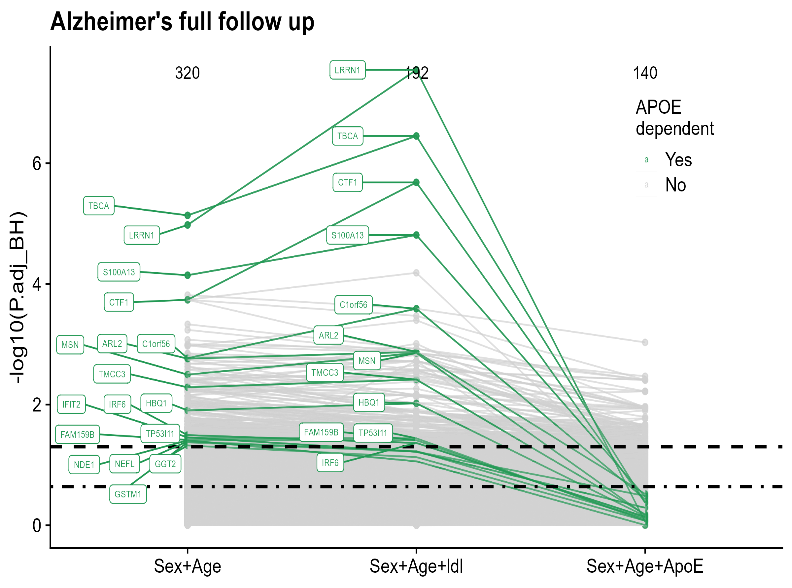

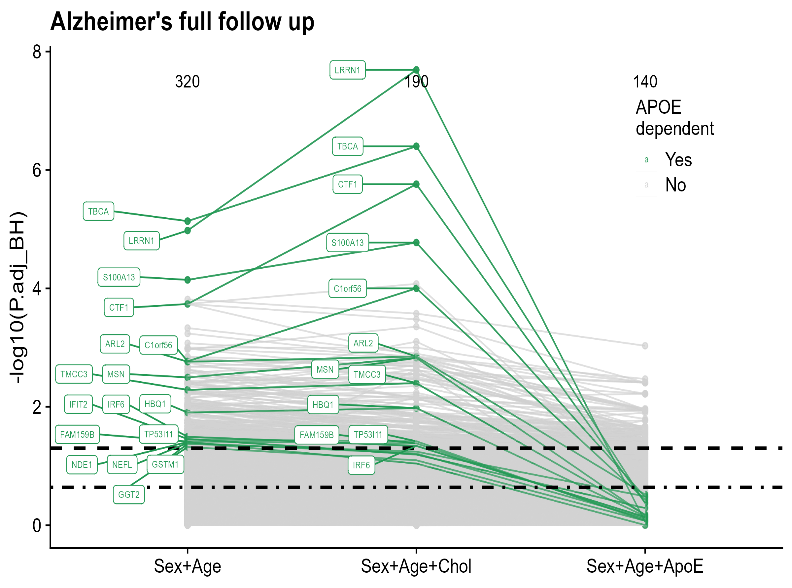

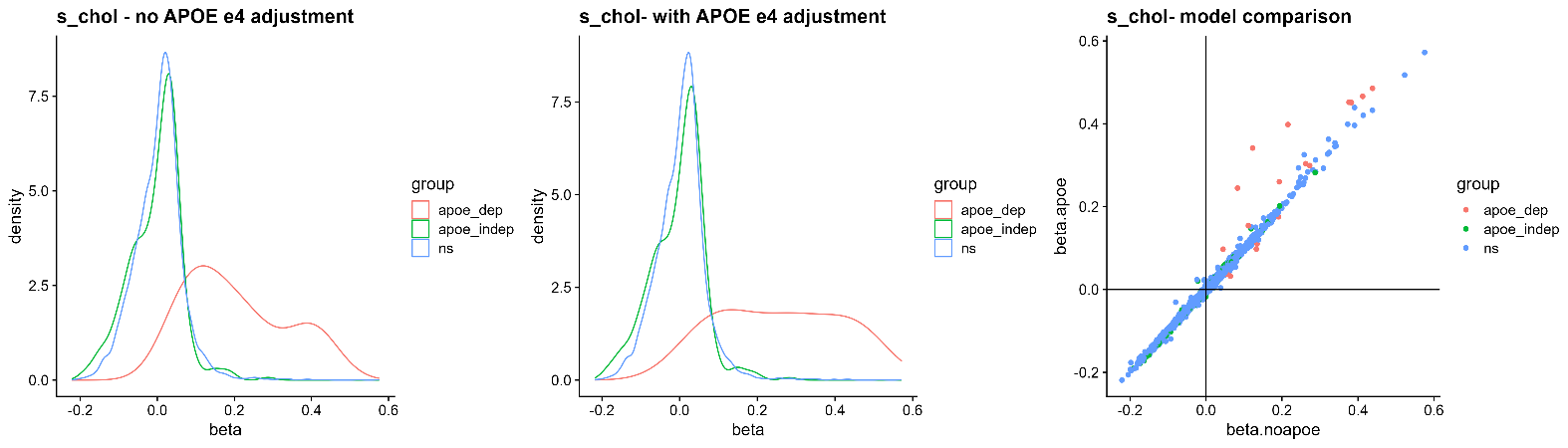

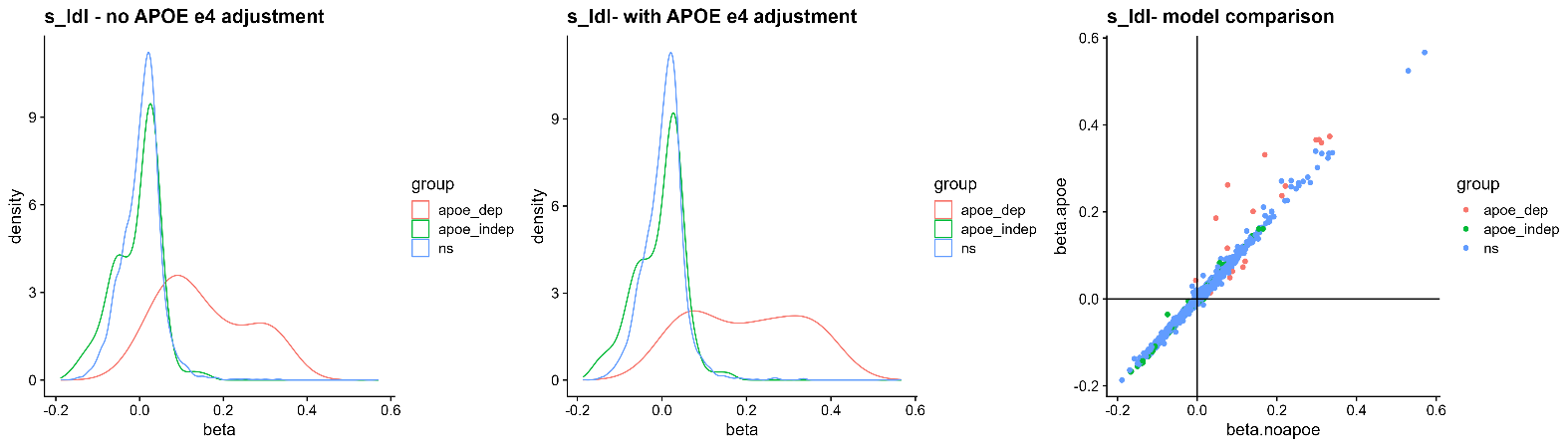

**A**

**B**

**C**

**D**

**E**
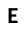

**F**
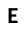

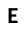

**G**
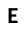

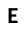

**H**
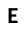

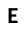

Supplementary Figure 6 – **A-B)** Spaghetti plots showing the statistical significance of protein associations with incident LOAD across three statistical models, adjusting for age and sex, and then additionally either serum **A)** total cholesterol or **B)** LDL cholesterol, and finally APOE e4 carrier status. The LOAD association of the 17 *APOE*-dependent proteins (green) is not attenuated when adjusting for total cholesterol or LDL but only when adjusting for APOE e4 carrier status. **C-H)** Protein associations with serum total cholesterol **(C-E)** and LDL cholesterol **(F-H)**. The density plots show the effect size for the associations between cholesterol levels and all measured proteins stratified by their AD association, adjusting for age and sex (**C** and **F**), and additionally *APOE-e4* carrier status (**D** and **G**). **E)** and **H)** Comparisons of the effect sizes for association with cholesterol levels in the models with (y-axis) and without (x-axis) *APOE-e4* adjustment shows that the effect size is often increased for the *APOE*-dependent proteins (red) after *APOE-e4* adjustment.

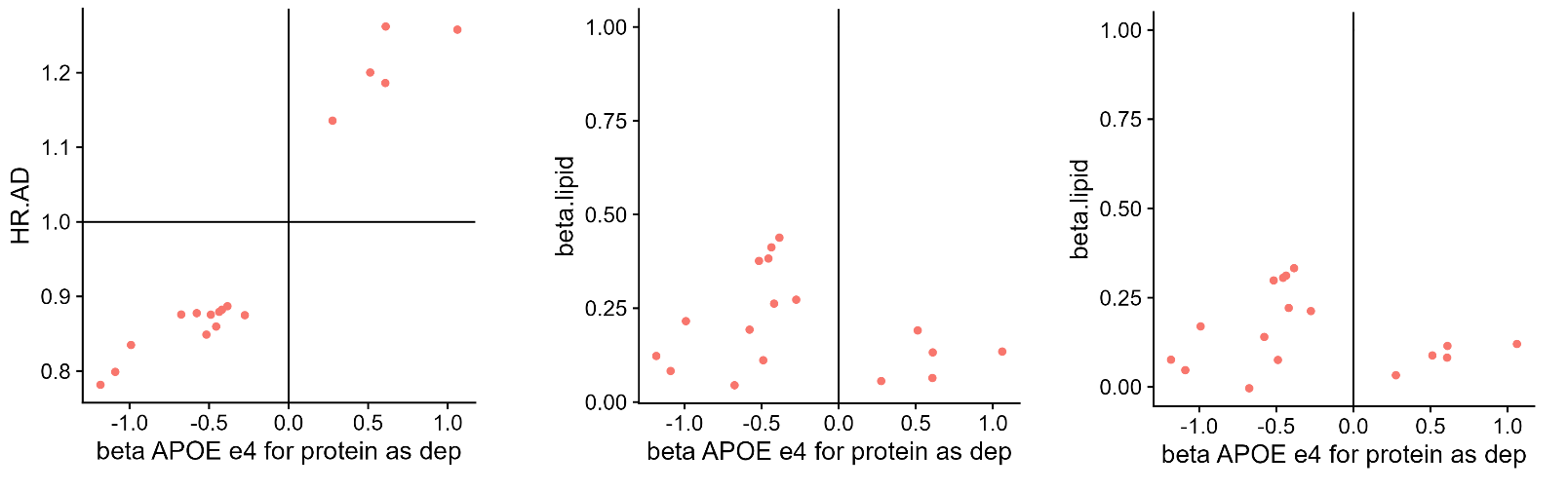

LOAD HR

TC Beta

LDL Beta

APOE-ε4 Beta

APOE-ε4 Beta

APOE-ε4 Beta

**C**
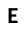

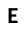

**D**
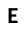

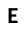

**E**
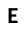

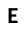

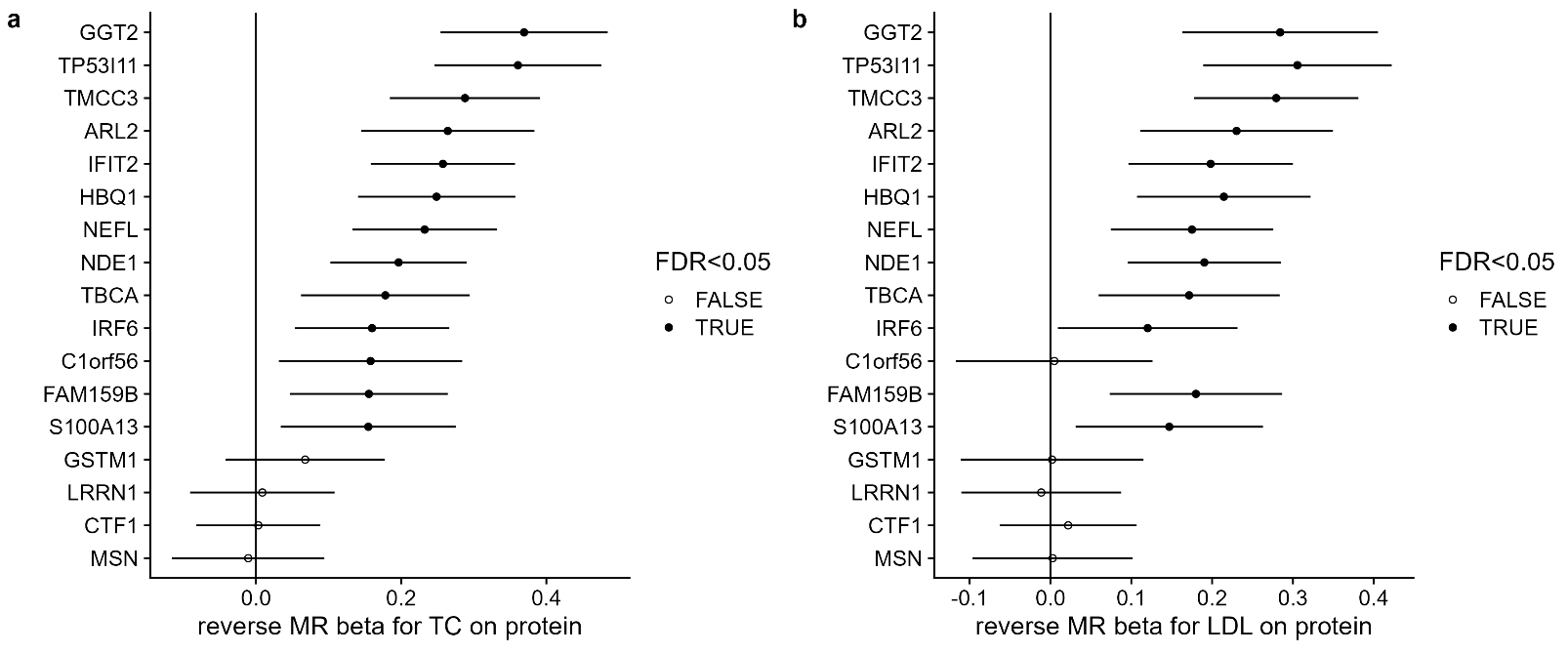

**A**
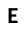

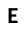

**B**
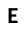

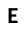

Supplementary Figure 7 – Reverse MR analysis (excluding the *APOE* locus) for the causal effect of **A)** total cholesterol and **B)** LDL cholesterol on serum levels of the 17 *APOE*-dependent proteins. **C-E)** For each protein, a comparison of the effect sizes for the association with the *APOE-ε4* genotype (x-axis) and **C)** incident LOAD, **D)** total cholesterol and **E)** LDL cholesterol (y-axis). TC, total cholesterol; LDL, low density lipoprotein cholesterol.

Supplementary Figure 8 – MR scatterplots for all proteins with more than one genetic instrument and with P<0.05 in the MR analysis for a causal effect of protein levels on AD. Each point represents a genetic instrument (SNP) and shows its effect on serum protein levels (x-axis) and AD (y-axis). The slope of the blue line indicates the inverse variance weighted MR effect.

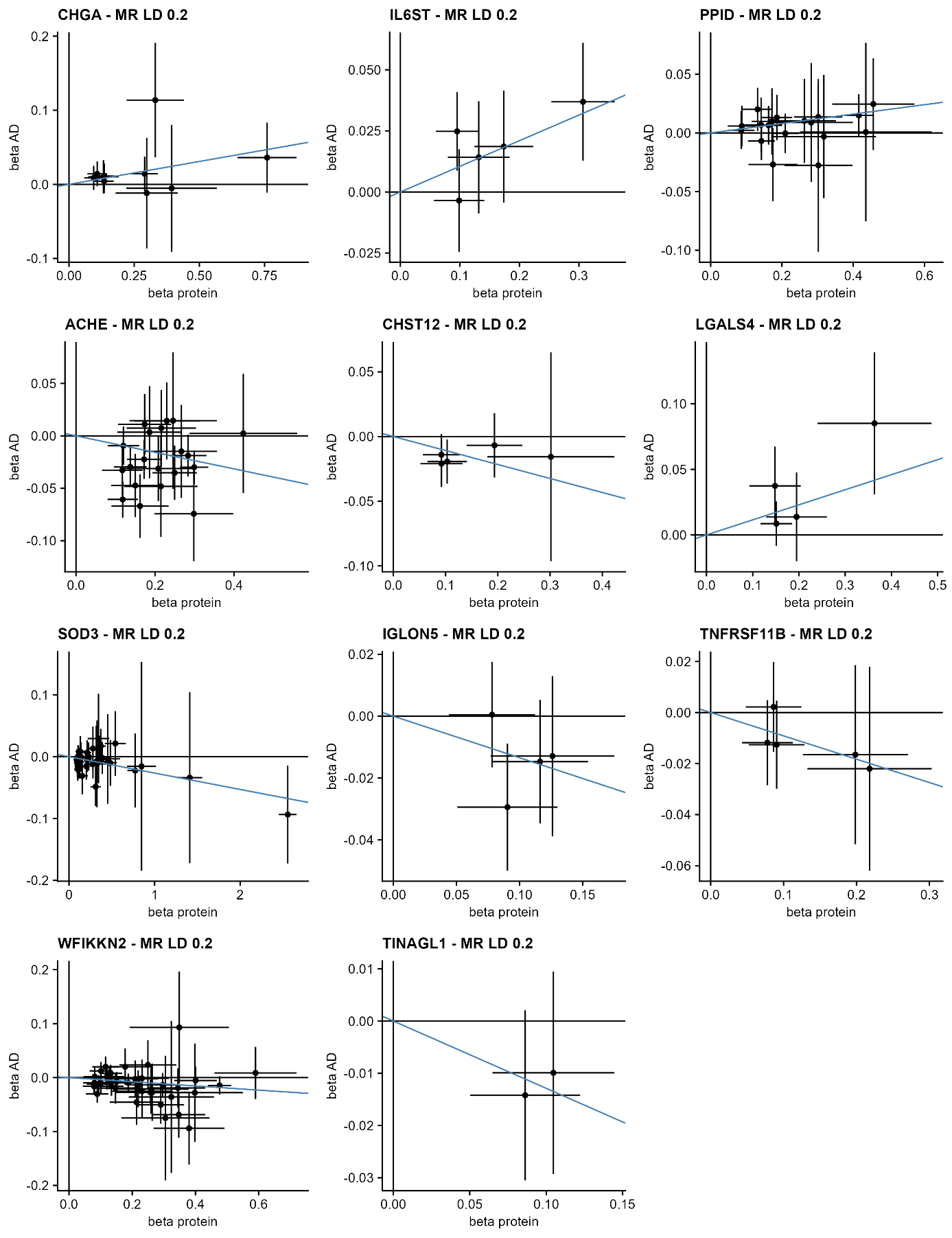

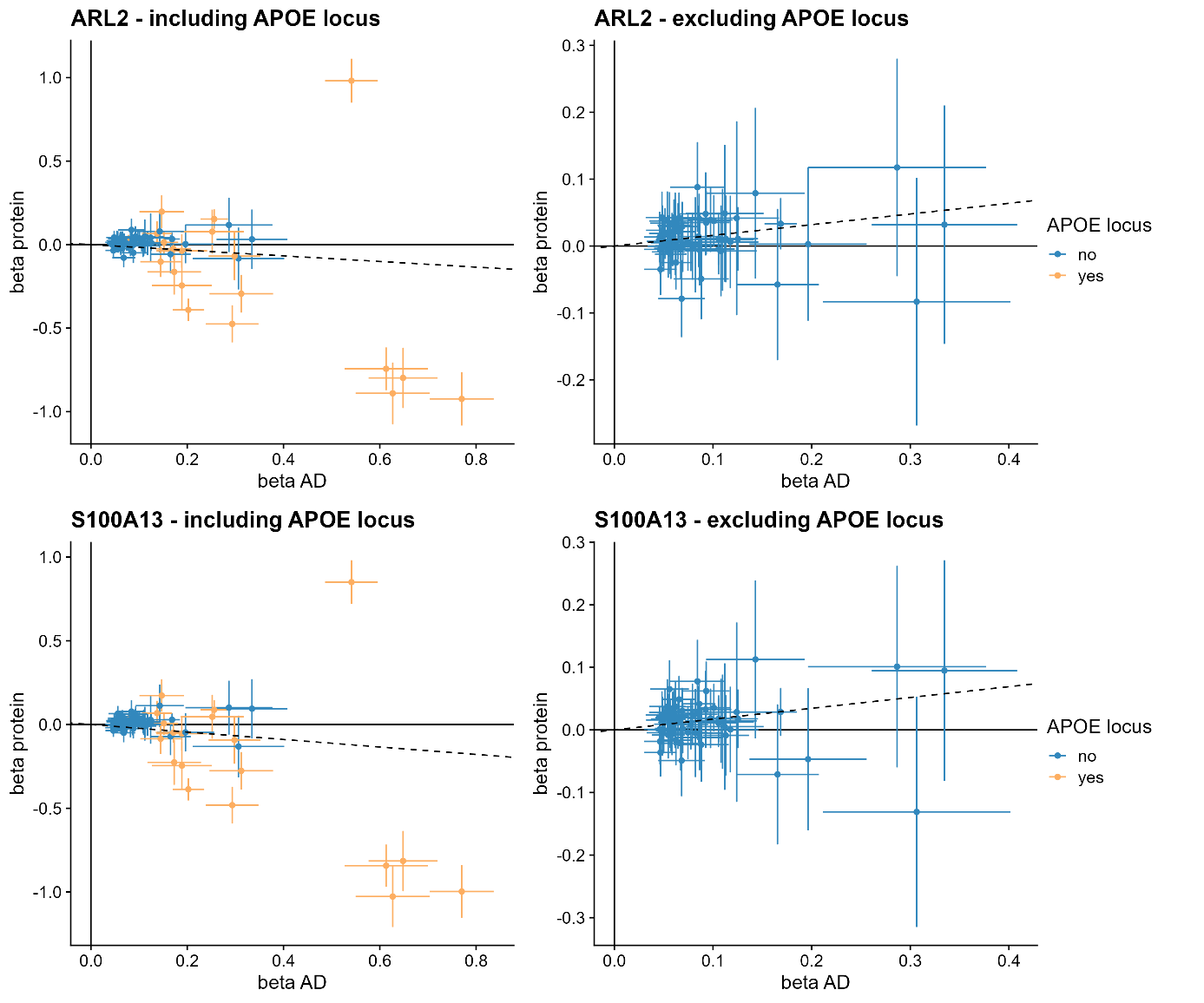

Supplementary Figure 9 – Example MR scatterplots from the reverse MR analysis for a causal effect of AD on serum protein levels, showing discordant direction of effect (inverse variance weighted method indicated by slope of dashed line) when including (left) or excluding (right) APOE genetic variants. Each point represents a genetic instrument (SNP) and shows its effect on AD (x-axis) and serum protein levels (y-axis).

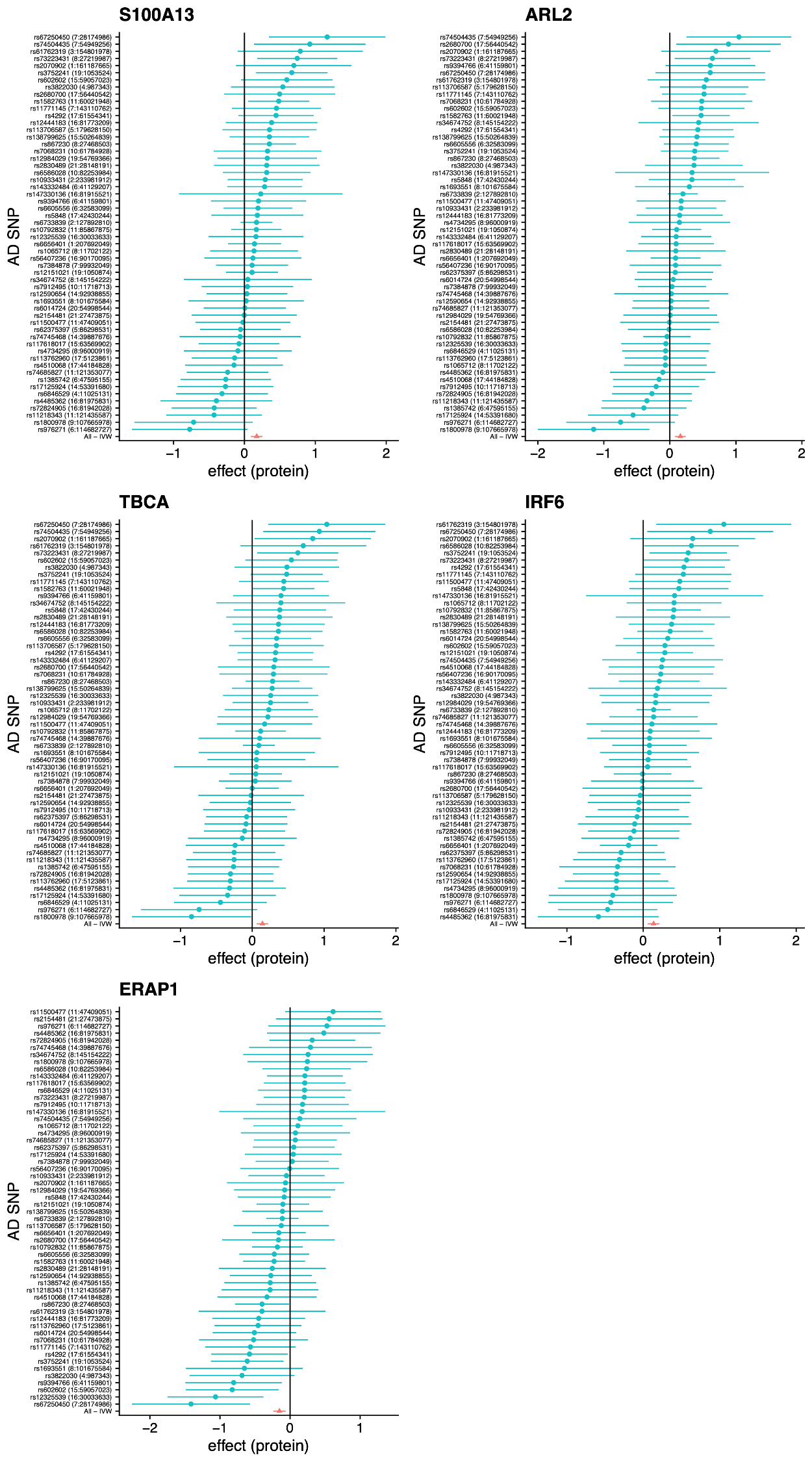

Supplementary Figure 10 – A forest plot showing the results for individual SNPs in the reverse MR analysis for a causal effect of AD on serum protein levels, together with the full multi-SNP IVW estimate (red). Plots are shown for the five proteins with FDR<0.1 in the primary reverse MR analysis, excluding variants from the *APOE* locus.

Supplementary Figure 11 – Comparison of the effects of *APOE-ε4* (left) versus AD, as evaluated through reverse MR analysis excluding *APOE* variants, (right) on serum protein levels in AGES and two additional cohorts. Results are shown for the four *APOE*-dependent proteins with opposite direction of effect in the two analyses. The AGES results are shown for the full cohort and two age strata.

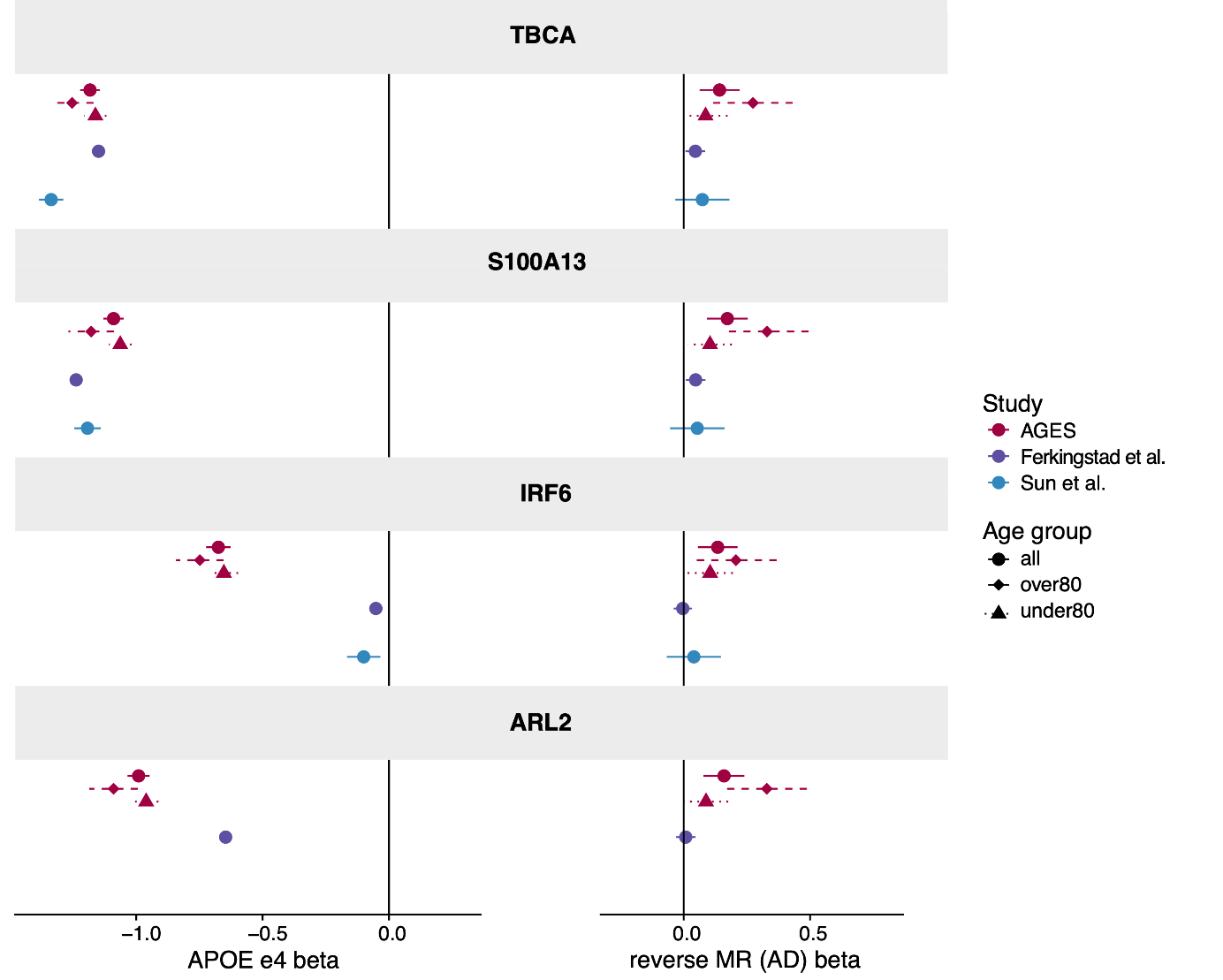

### Supplementary references

1.	Li, T. *et al.* A scored human protein–protein interaction network to catalyze genomic interpretation. *Nature Methods 2016 14:1* **14**, 61–64 (2016).

2.	Postmus, I. *et al.* Pharmacogenetic meta-analysis of genome-wide association studies of LDL cholesterol response to statins. *Nat Commun* **5**, (2014).

3.	Menzel, H., Kladetzky, R. & Assmann, G. Apolipoprotein E polymorphism and coronary artery disease. *Arteriosclerosis* **3**, 310–315 (1983).

4.	Stengård, J. H. *et al.* Apolipoprotein E Polymorphism Predicts Death From Coronary Heart Disease in a Longitudinal Study of Elderly Finnish Men. *Circulation* **91**, 265–269 (1995).
